# Modeling and Preparedness: The Transmission Dynamics of COVID-19 Outbreak in Provinces of Ecuador

**DOI:** 10.1101/2020.07.09.20150078

**Authors:** Carlos Bustamante-Orellana, Jordy Cevallos-Chavez, Cesar Montalvo-Clavijo, Jeff Sullivan, Edwin Michael, Anuj Mubayi

## Abstract

Coronavirus disease 2019 (COVID-19), a novel infectious disease first identified in December 2019 in the city of Wuhan of China’s Hubei province, is caused by severe acute respiratory syndrome coronavirus 2 (SARS-CoV-2). The disease has become a pandemic in just a few months and spread globally with more than 2.89 million cases and 203,000 deaths across 185 countries, as of April 26th, 2020. Ecuador has reported one of the highest rates of COVID-19 in Latin America, with more than 10K cases and 500 deaths in a country of approximately 17 million people. The dynamics of the outbreak is being observed quite different in different provinces of Ecuador with high reported prevalence in some low population density provinces. In this study, we aim to understand variations in outbreaks between provinces and provide assistance in essential preparedness planning in order to respond effectively to ongoing COVID-19 outbreak. The study estimated the critical level of quarantine rate along with corresponding leakage in order to avoid overwhelming the local health care system. The results suggest that provinces with high population density can avoid a large disease burden provided they initiate early and stricter quarantine measures even under low isolation rate. To best of our knowledge, this study is first from the region to determine which provinces will need much preparation for current outbreak in fall and which might need more help.

## 1 Introduction

The World Health Organization declared the Coronavirus outbreak (Covid-19) as a public health emergency of international concern on the 30th of January 2020. By 11^*th*^of March, the Covid-19 spread across globe affecting around 114 countries and killing more than 4600 individuals. It was immediately elevated officially to the category of pandemic by WHO. After more than five months since the first case was reported in Wuhan, China, the cases reached more than 10 million and half million deaths across all the countries, as of June 30, 2020 [1, 2].

In early February, the World Health Organization reported that the virus causing the disease, is a novel strain of coronavirus (nCoV-2019), [3]. Coronaviruses are a large family of viruses that usually cause respiratory diseases ranging from the common cold to severe respiratory illnesses such as SARS (2003) and MERS (2012), [4]. In mid February, the International Committee on Taxonomy of Viruses (ICTV) named this novel virus as Severe Acute Respiratory Syndrome Coronavirus 2 (SARS-CoV-2) since it is caused by a mutation of the coronavirus related with SARS; and the disease name was designated by the WHO as COVID-19, short for coronavirus disease 2019, [5, 3].

The Coronavirus is assumed to be transmitted via droplets and fomites during close unprotected contacts with infected individuals or with surfaces that may have been exposed. The new coronavirus is believed to affect the lungs, and other the respiratory system. Some of the most common (but distinct from other similar diseases) symptoms of COVID-19 include dry cough with high fever, shortness of breath, chills and loss of taste or smell [3, 6, 7]. It has been seen that around 80% of infected people either experience a mild form of disease or are asymptomatics and around 5% become critically ill. Prepared health care systems may be able to handle some of the critically ill patients, however, the problem remains that even the most sophisticated healthcare systems may get overwhelmed by high number of hospitalizations if proper social distancing measures are not timely implemented.

On the other hand, non-pharmaceutical interventions, such as quarantine, isolation and the use of masks for general population, have been implemented in many countries to reduce the number of secondary infections [6, 8], however, a large variations is observed in initiation and magnitude level of implemented interventions. Economic stress due to social distancing measures are making it difficult to put strict guidelines on non-pharmaceutical interventions. When and at what level certain types of non-pharmaceutical interventions need to be implemented, remains a big question in controlling the current COVID-19 pandemic, especially in resource-limited regions.

In Ecuador, the first positive case of Covid-19 was detected on February 29th, 2020. The infected person was a 70 years old woman coming from Spain, who landed in Ecuador on February 14th. The dramatic increase in the number of cases and the concentration of infections in Guayaquil, the largest city in Ecuador, has described this city as the center of the pandemic in Latin America. The ministry of Health confirmed the death of patient zero on March 13th, 2020, after being in intensive care unit. The number of cases has substantially increased after the first case, amidst a health system that has collapsed due to the growing contagion in some parts. By the end of April, Ecuador registered more than 20,000 confirmed cases of Covid-19 with 1000 deaths. Since the 12th of March, authorities have closed schools, large events, and cancelled international flights. From the 17th of March, the government restricted services of public transit system within the country. On the other hand, the spreading of the virus has impacted the different provinces of the country in different ways, with early data is showing the provinces with higher population density are the most affected ones.

In this study, we develop two types of models (i) a simple model capturing initial growth of an epidemic and (ii) a complex transmission dynamics model that studies the role of various epidemiological mechanisms in driving an outbreak. Using these models, we assessed the impact of quarantine, and isolation strategies on the the number of new COVID-19 related hospitalizations and cumulative deaths in Ecuador and eight of its provinces. The goals of this study are to: (i) estimate initial growth rate for each of the eight most affected provinces, (ii) compute the reproduction number for the provinces and the Ecuador, (iii) evaluate the impact of quarantine and isolation policies on the dynamics of the disease under limited healthcare resources, and (iv) compare and contrast differences in outbreak in 8 provinces with respective constraints in resources.

## 2 Methods

### 2.1 Data Sources

#### Description of Time Series Data

The data about positive confirmed cases and deaths of Covid-19 in Ecuador has been obtained from two main sources: the Ministerio de Salud Pu’blica del Ecuador [9] and Servicio Nacional de Gesti’on de Riesgos y Emergencias [10]. These two public data sources started to publish daily situation reports about cases of Covid-19 in Ecuador since March 13rd, 2020. The data provided by these sources consisted of information about the number of confirmed cases, suspected cases, deaths, discarded cases, and recovered cases for each province of the country. These information were gathered from each daily reports and organized for further analysis.

#### Description of Hospitalization Data

The information on hospital resources for each province was obtained from the Instituto Nacional de Estad’isticas y Censos [11] from Ecuador. This source provided information about the number of hospital beds available in each province in 2018. The occupancy rate for Ecuador was also obtained from WHO [12] and used together with the number of beds available in each province, to estimate the number of available beds for COVID-19 patients in each province.

### 2.2 Model Description

#### 2.2.1 Initial Growth Models

The growth rate of an epidemic is a key measure of the severity at which the epidemic can impact a population, and it is closely related to the basic reproduction number (*ℛ*_0_) [13]. At the beginning of an epidemic, the model that describes an exponential growth rate in the number of confirmed cases of infected people is:

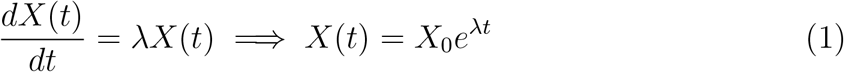

where *X*(*t*) is the number of cases at time *t, X*_0_ is the number of initial cases, and, λ is the rate of growth.

The initial cumulative number of cases, *Y* (*t*), often has been observed to grow slower than exponential in scenarios that include restricted contact among individuals [14]. Hence, we also considered a simple generalized model that relaxes the exponential growth rate as assumption:

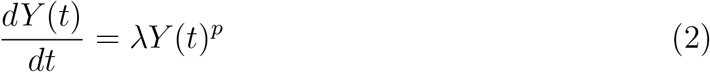

where, *Y* (*t*) is the cumulative number of cases at time t,, λ is a positive parameter denoting the growth rate (1/time), and *p* ∈ [0, 1] is a deceleration of growth parameter. If *p* = 0, this equation describes constant incidence over time and the cumulative number of cases grows linearly while *p* = 1 models exponential growth dynamics (i.e., Malthus equation). Intermediate values of p between 0 and 1 describe sub-exponential (e.g., polynomial) growth patterns [15]. For sub-exponential growth (i.e., 0 < p < 1) the solution of this equation is given by the following polynomial of degree *m*:

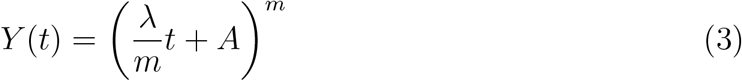

where, *m* is a positive integer, and the deceleration of growth parameter is given by *p* = 1 − 1/*m*, and 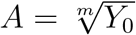.

#### 2.2.2 COVID-19 Dynamical Model

We considered a SEAIR (Susceptible-Exposed-Asymptomatic-Infectious Symptomatic-Recovered) type epidemic model with additional features such as closed population (i.e., *N* is constant) and presence of interventions such as quarantine (*Q*_1_, *Q*_2_, and *Q*_3_), home isolation (*Q*_4_), hospitalization or hospital-isolation (*H*) and Intensive care unit (ICU, *C*) classes. Infection can lead to transition of susceptible individuals to the latent (infected but not infectious and not showing symptoms) class. Latent individuals can progress to asymptomatic infectious (*A*) class. The population from susceptible, latent and asymptomatic classes can self quarantine to *Q*_1_, *Q*_2_ and *Q*_3_ classes, respectively. Symptomatic individuals (*I*) can directly go to hospitals (*H*), recover (*R*), get isolated (*Q*_4_), or die due to the disease (*D*). We assume that the quarantine is not perfect (that is, leakage in quarantine) and the proportion *l*_1_ of the quarantined population will not follow it strictly. Moreover, leaked quarantined individuals may be infectious by a factor of *ϵ*_1_. We also consider the proportion *l*_2_ and *l*_3_ of isolated individuals in *Q*_4_ and *H* classes, respectively, may not be strictly isolated (leading to additional infection by them). This may be due to limited equipment and resources or economic burden due to unable to work. The model assumes that the infectivity of individuals in *H* class may lower than I by a factor of *ϵ*_2_. It is further assumed that patients in ICU do not mix with other individuals in the population due to their inability to leaving hospital as a result of severe health condition. The model assumes that people from Intensive Care Unit (ICU) only recover after being in observation, i.e. people go out from ICU to recovered via hospitalization. The flowchart of the compartmental model is given in Figure 1 and its variables and parameters are defined in Table 1 and Table 2, respectively.

**Table 1:**
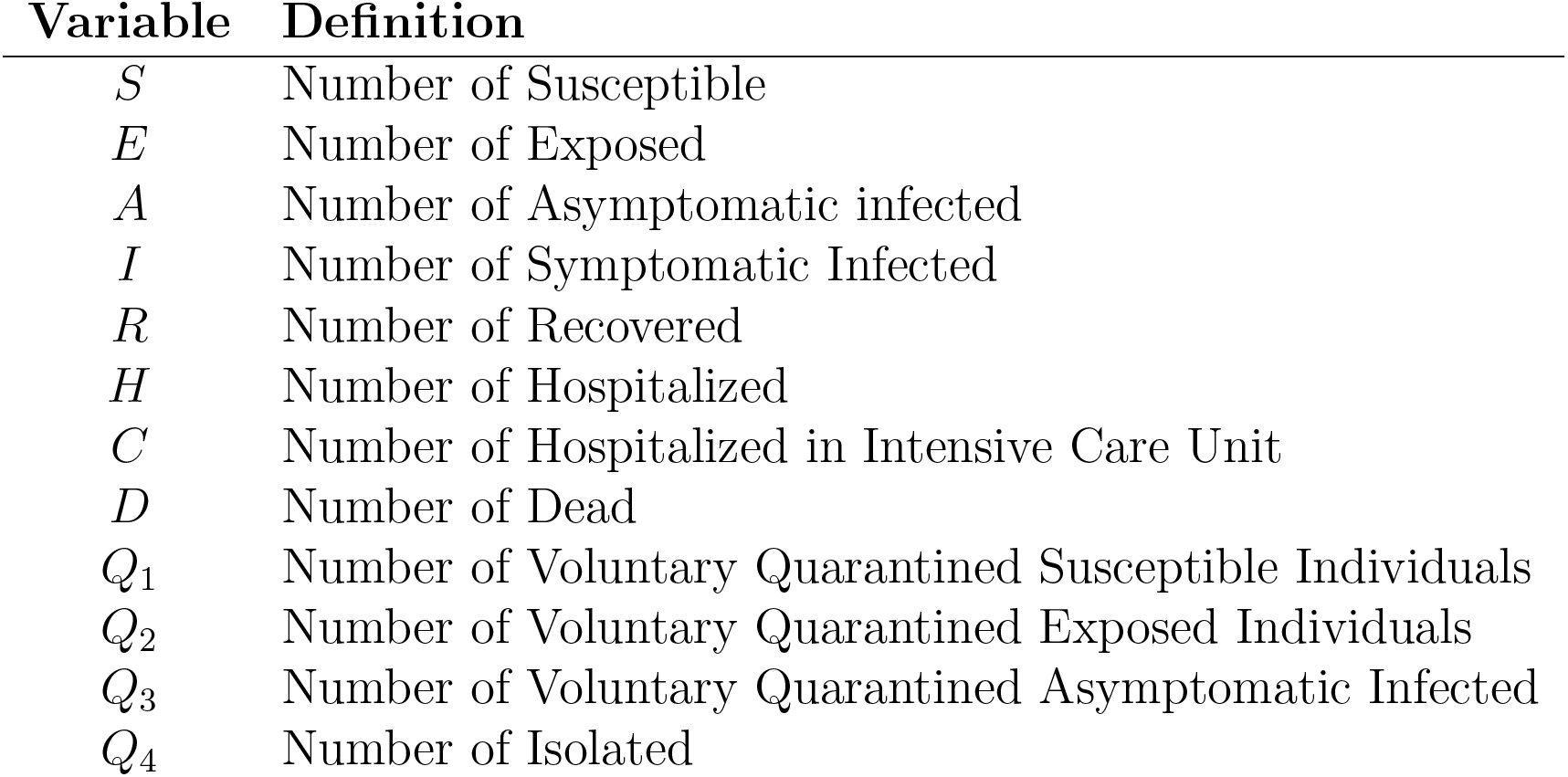
Model variables and their definition

| Variable | Definition |
| --- | --- |
| $S$ | Number of Susceptible |
| $E$ | Number of Exposed |
| $A$ | Number of Asymptomatic infected |
| $I$ | Number of Symptomatic Infected |
| $R$ | Number of Recovered |
| $H$ | Number of Hospitalized |
| $C$ | Number of Hospitalized in Intensive Care Unit |
| $D$ | Number of Dead |
| $Q_1$ | Number of Voluntary Quarantined Susceptible Individuals |
| $Q_2$ | Number of Voluntary Quarantined Exposed Individuals |
| $Q_3$ | Number of Voluntary Quarantined Asymptomatic Infected |
| $Q_4$ | Number of Isolated |

**Table 2:**
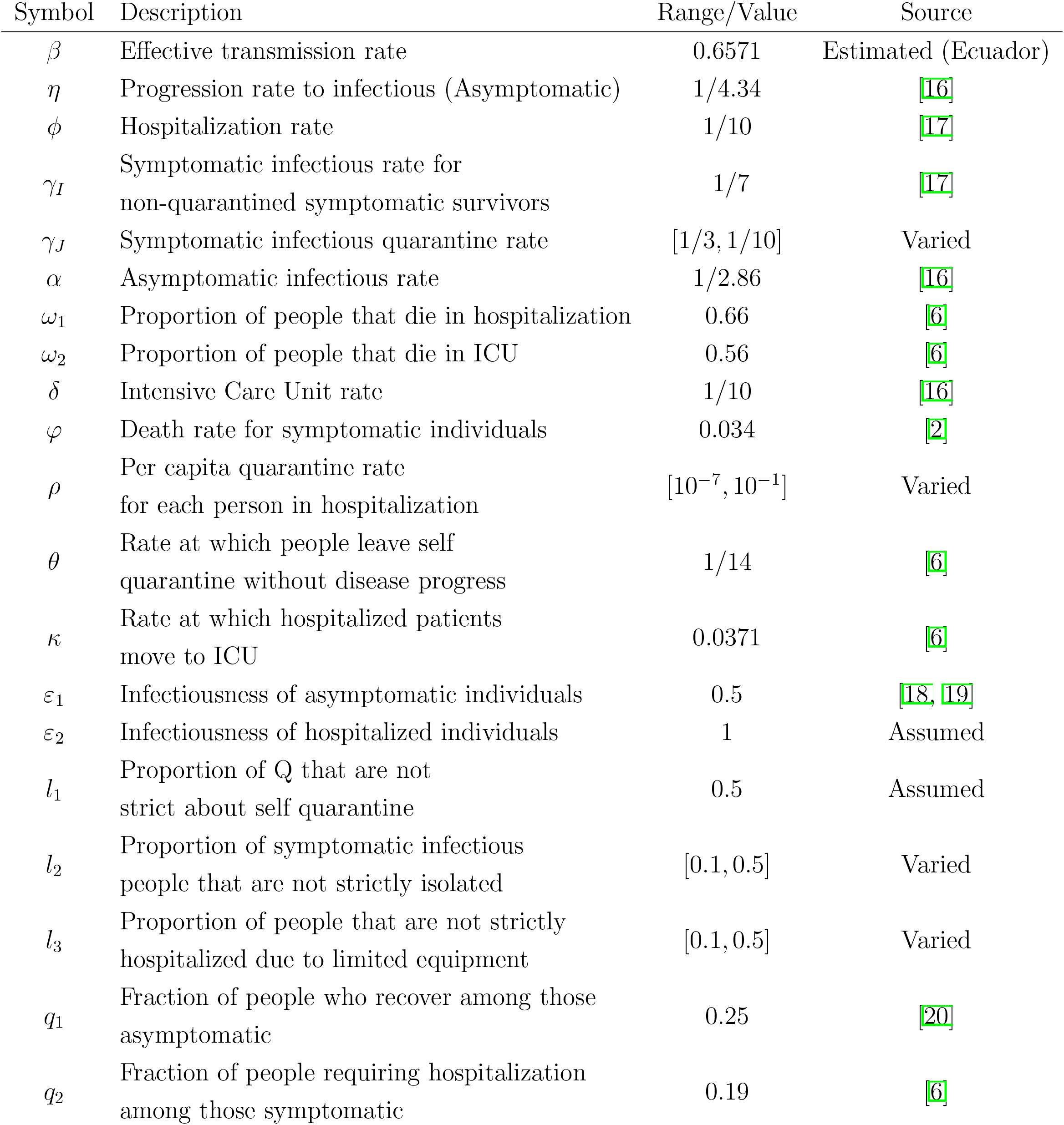
Description of parameter values and respective values obtained from literature

**Figure 1:**
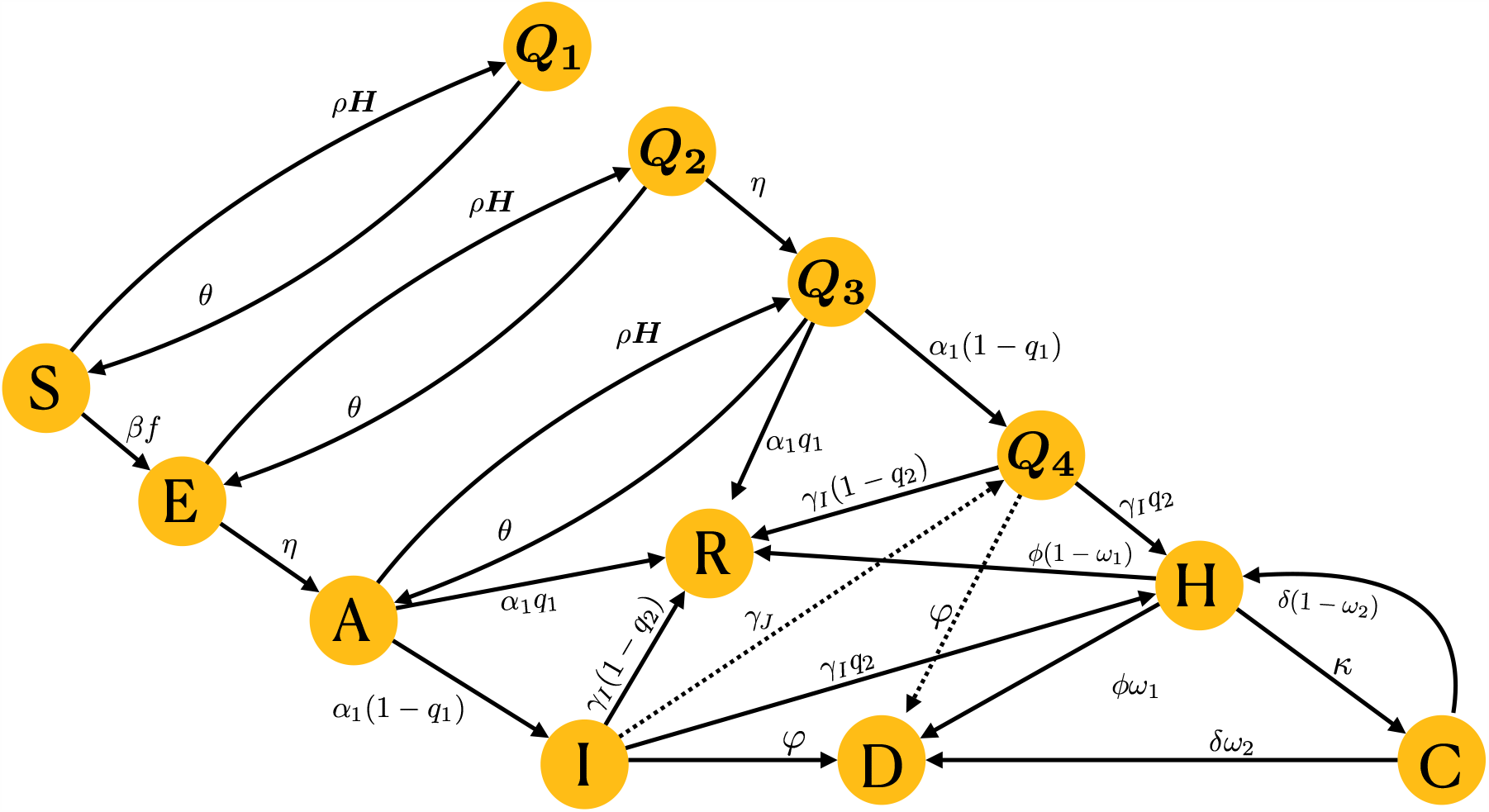
Schematic flow diagram for our model. The model consists of twelve subpopulations and their definitions are given in Table 1.

The force of infection, f, is a nonlinear term given by

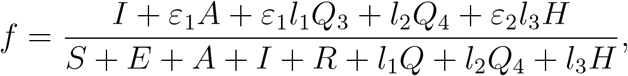

where *Q* = *Q*_1_ +*Q*_2_ +*Q*_3_. Moreover, the parameters *τ* and *ω*_1_ capture limited resources (limited number of ICU beds, *C*_0_) conditions in healthcare facilities and hence,

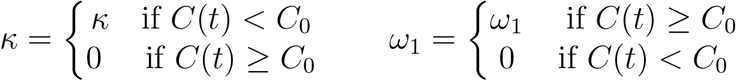

The parameter *τ* represents the rate at which hospitalized people are moved to ICU, so it is zero if there are no ICU beds available; and the parameter *ω*_1_ is the proportion of people that die in hospitalization, so it is zero if there are ICU beds available because they would die in ICU, not in normal hospitalization. Our COVID-19 epidemiological model is given by the non-linear system of differential equations given in Appendix B.2.

## 3 Analysis

### 3.1 The Reproduction Number and Final Epidemic Size

The local stability of the Disease Free Equilibrium (DFE) is analyzed by using the Next Generation Operator method [21]. We could obtain that the *ℛ*_0_ for the model without any control, i.e. when voluntary quarantine is not applied, i.e. *ρ* = *θ* = 0, when Mandatory quarantined is not applied, *q*_4_ = *q*_3_ = 1, and when hospitalization and ICU are not available neither, i.e *ϕ* = 0 and *κ* = 0.

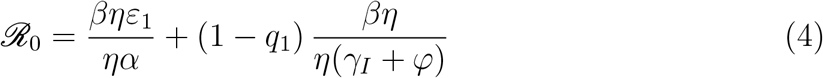

Furthermore, the final epidemic size for the baseline model, computed in Appendix C.3, is given implicitly as follows

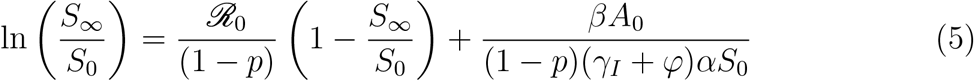

Moreover, the control reproduction number, denoted by ℛ_*c*_, orthecompletemodel computed in Apendix C.2, will be

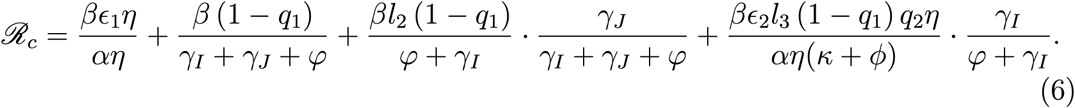

### 3.2 The Initial Exponential Growth Rate from the Dynamic Model

At the beginning of an epidemic, the number of cases tend to grow exponentially if no strict measurements are taken. This type of growing in the number of Covid-19 cases happened in many provinces of Ecuador, especially Guayas, which is the most affected by the virus. Therefore, computing the initial exponential growth rate for the number of cases in each province of Ecuador can help us model the dynamics of the disease within such province.

In order to determine the initial exponential growth rate from a model, we linearize the model about the disease-free equilibrium (DFE). For this particular study, we consider a reduced version of the model sketched in Figure 1, where the only interventions are hospitalization and ICU. So, this reduced model would only have the Susceptible (S), Exposed (E), asymptomatic infected (A), symptomatic Infected (I), Recovered (R), Hospitalized (H), Hospitalized in Intensive Care Unit (C), and Death (D) compartments. The DFE of this model is: (*S, E, A, I, H, C, R, D*)=(*N*, 0, 0, 0, 0, 0, 0). We linearize the reduced model about the DFE, the equations for E, A, I, H, and C are decoupled, and become:

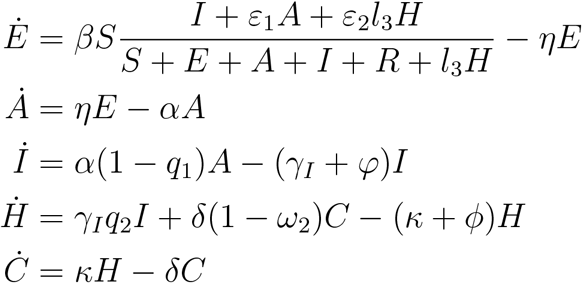

Note that the Jacobian matrix

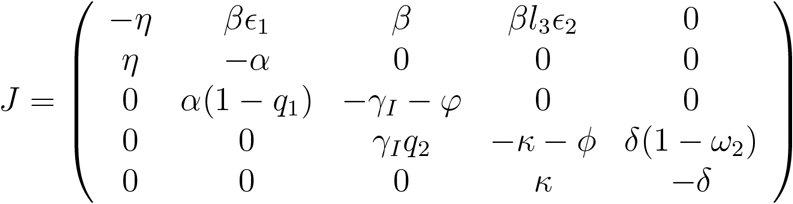

has the following characteristic equation:

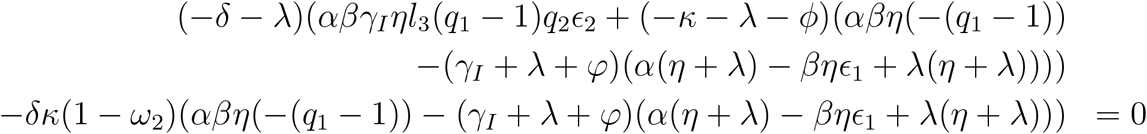

where, λ are the eigenvalues of the Jacobian evaluated at the DFE. The initial exponential growth rate is the largest root, λ of the previous fifth degree equation. The initial exponential growth rate can be measured experimentally, and if the measured value is, λ, then from the previous equation, we obtain:

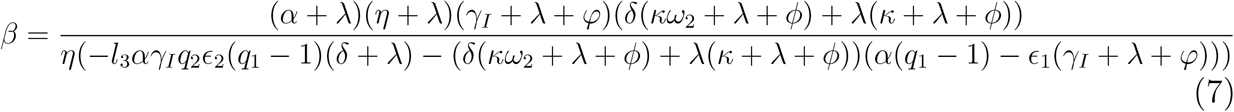

So, we can estimate *β* from the initial exponential growth rate. Besides, as *ℛ*_*c*_ depends on *β*, we can also estimate *ℛ*_*c*_ from the initial exponential growth rate by plugin the equation 7 into the formula for *ℛ*_*c*_.

We obtained, λ for the confirmed Covid-19 cases and deaths using data provided by official sources of Ecuador [9, 10]. The obtained data had the total confirmed cases and deaths of Covid-19 for the 24 provinces of Ecuador from March 13^*rd*^ until April 16^*th*^, 2020.

### 4 Results

### 4.1 Exponential Epidemic Growth Model Results

We fitted the model in Equation 8 to the cumulative data of confirmed cases in 8 provinces of Ecuador and the whole country, and the obtained results are presented in Table 3. The relationship between the variables was measured using the adjusted R squared (*SQR*) and a 95% confidence interval was computed for the fitted exponential growth rate (λ). The estimated first case was also obtained, using the parameters fitted to the data, and compared with the first reported case.

**Table 3:** Exponential Fitting results for total confirmed cases in Ecuador and 8 of its most affected provinces.

| Province / Country | Exponential Fitting Confirmed Cases | | | Confidence Interval for $\lambda$ ($CI = 95\%$) | | First Reported Case | Estimated First Case |
| --- | --- | --- | --- | --- | --- | --- | --- |
| | $I_0$ | $\lambda$ | SQR | 2.5% | 97.5% | | |
| Guayas | 47 | 0.1636 | 0.8220 | 0.1371 | 0.1900 | 02/29/20 | 02/17/2020 |
| Los Rios | 9 | 0.1097 | 0.9634 | 0.1022 | 0.1171 | 03/10/20 | 02/21/2020 |
| Pichincha | 8 | 0.1514 | 0.8987 | 0.1337 | 0.1691 | 03/12/20 | 02/27/2020 |
| Sucumbios | 1 | 0.1194 | 0.8101 | 0.0993 | 0.1395 | 03/13/20 | 03/10/2020 |
| Azuay | 3 | 0.1398 | 0.8240 | 0.1169 | 0.1626 | 03/17/20 | 03/04/2020 |
| Manabi | 3 | 0.1398 | 0.8405 | 0.1175 | 0.1620 | 03/18/20 | 03/04/2020 |
| El Oro | 1 | 0.1697 | 0.8935 | 0.1478 | 0.1915 | 03/20/20 | 03/12/2020 |
| Santo Domingo | 1 | 0.1322 | 0.8959 | 0.1144 | 0.1500 | 03/21/20 | 03/11/2020 |
| Ecuador | 78 | 0.1565 | 0.8636 | 0.1349 | 0.1782 | 02/29/20 | 02/13/2020 |

The same above process was repeated to fit the confirmed cumulative deaths to the Model Equation 8, obtaining the results presented in Table 10. Note, that Sucumbios province was not considered due to not enough availability of data for fitting.

#### 4.2 Generalized Epidemic Growth Model Results

We fitted the Model Equation 9 to the cumulative confirmed cases of Covid-19 in Ecuador and obtained the best values for each parameter of the model (see Table 4). The parameter *p*, which is used to decelerate the growth of the epidemic, has also been estimated. *The values for this parameter suggest that in most of the provinces of Ecuador, and in the whole country*, the growth rate of the cumulative number of cases is not exponential, but quadratic. It is important to mention that the residual standard error (RSE) is considerably bigger for Ecuador and Guayas than for any of the other provinces. This difference may be caused by the arrival of a large amount of testing kits for Covid-19 on a particular day, which generated a considerable difference in the amount of positive tests processed for that day than the average tests that were being process on a daily basis. Although all of the provinces may have been affected by this change in the number of tests processed by day, Guayas is the province that concentrates most of the cases of Covid-19 in Ecuador, and therefore most of the extra tests processed in such an abnormal day were from that province.

**Table 4:** Generalized growth fitting (Model 9) results for the cumulative confirmed cases of Covid-19 in Ecuador and 8 of its most affected provinces

| Province /<br>Country | $\lambda$ | $p$ | $X_0$ | RSE |
| --- | --- | --- | --- | --- |
| El Oro | 0.39 | 0.72 | 0.21 | 9.29 |
| Manabi | 0.40 | 0.70 | 1.22 | 13.09 |
| Santo Domingo | 0.26 | 0.69 | 1.00 | 2.95 |
| Guayas | 2.62 | 0.57 | 13.26 | 355.80 |
| Los Rios | 0.93 | 0.49 | 1.00 | 11.52 |
| Pichincha | 1.31 | 0.53 | 1.00 | 20.15 |
| Azuay | 0.71 | 0.52 | 1.00 | 9.45 |
| Sucumbios | 0.78 | 0.13 | 1.00 | 4.62 |
| Ecuador | 3.72 | 0.55 | 5.59 | 405.60 |

The cumulative COVID-19 related deaths data were also fitted to the Model Equation 9 (see Table 5). In this case, the provinces Los Rios, Sucumbíos and Azuay were not considered because the mortality data available for these provinces were not statistically sufficient to perform a proper fitting.

**Table 5:** Generalized growth fitting (Model 9) results for the cumulative deaths of Covid-19 in Ecuador and 5 of its most affected provinces

| Province /<br>Country | $\lambda$ | $p$ | $X_0$ | RSE |
| --- | --- | --- | --- | --- |
| Manabi | 0.10 | 1.00 | 1.00 | 5.87 |
| Pichincha | 0.10 | 1.00 | 1.00 | 3.06 |
| El Oro | 0.11 | 0.99 | 1.00 | 2.39 |
| Santo Domingo | 0.10 | 0.58 | 1.00 | 2.31 |
| Guayas | 0.68 | 0.53 | 1.00 | 14.79 |
| Ecuador | 0.58 | 0.66 | 1.00 | 15.17 |

##### 4.3 The Dynamic Model-Based Numerical Results

We ran simulations for the Covid-19 model presented in this study using the parameter values from Table 2, and varying the values of the four unknown parameters (specified as varied in Table 2: *ρ, γ*_*J*_, *l*_2_, *l*_2_, and *l*_3_). The procedure was the following: one of the parameters was varied in a range and for each value of such parameter, fifty different values of the other three parameters were generated, and therefore, the model was run fifty times for each value of the parameter being varied. Then, the mean of the different results provided by those 50 simulations was computed and portrayed (see Figures 2-5).

**Figure 2:**
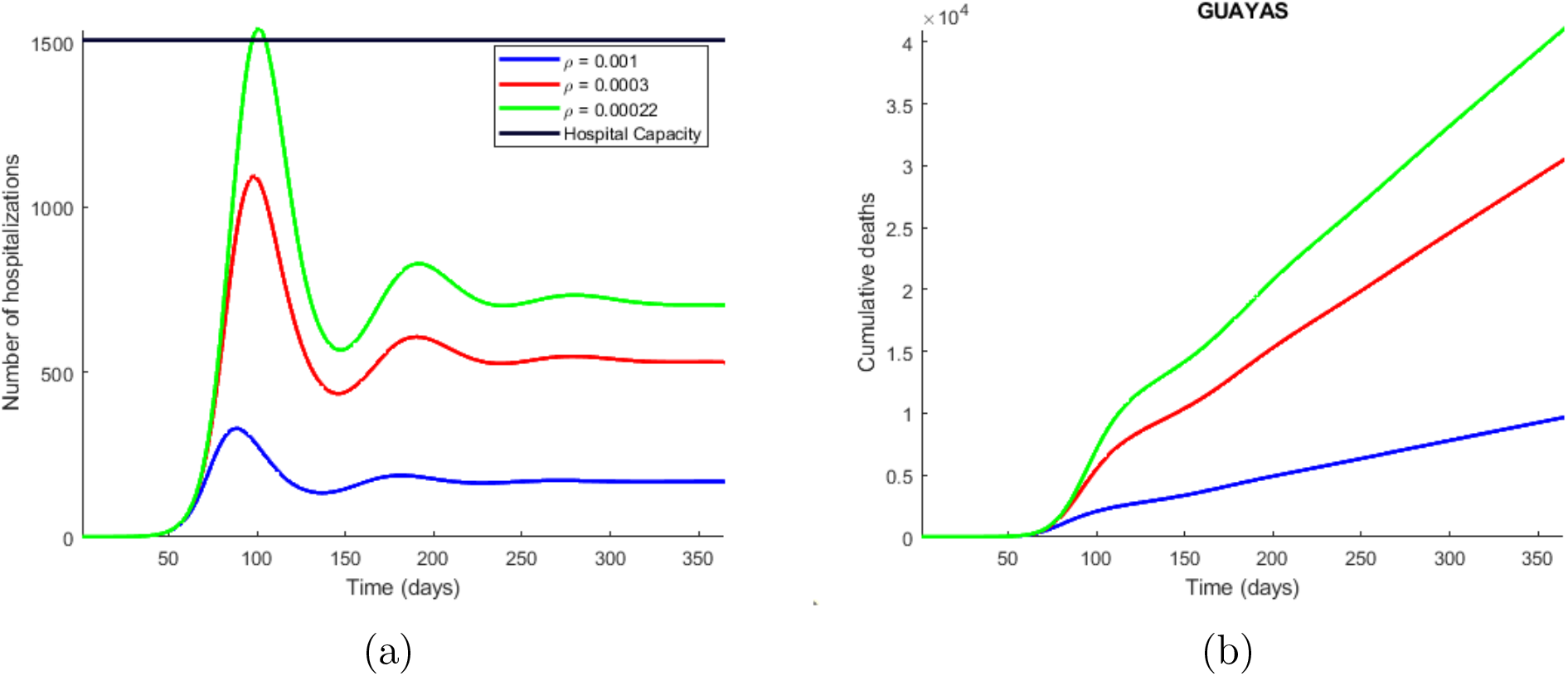
Time series plot of the number of hospitalizations (a), and cumulative deaths (b) predicted by the model. The simulations were done for Guayas province with the parameters values presented in Table 2 and varying the quarantine rate (*ρ*) between (10 of each 10000 susceptible individuals go to quarantine) and 0.0002. The legend specifies the values for the quarantine rates that produce the corresponding curves, considering the explained meaning of such values. The horizontal line represents the hospitals capacity in the province.

Estimations for the effective transmission rate (*β*) and control reproduction number (*ℛ* _*c*_) were obtained (see Table 6) using the formulas 7 and 6, respectively. As there were unknown parameters (*ρ, γ*_*J*_, *ϵ*_2_, *l*_2_), the estimations obtained for *β* and *ℛ* _*c*_ are actually the mean of 50 different values obtained from sampling 50 different values for the unknown parameters. The ranges for each unknown parameter are presented in Table 2.

**Table 6:** Estimations of effective transmission rate (*β*) and control reproduction number (*ℛ*_*c*_) for Ecuador and eight of its provinces.

| Province / Country | Estimated $\beta$ | Confidence Interval for $\beta$ ($CI = 95\%$) | | Estimated $R_c$ | Confidence Interval for $R_c$ ($CI = 95\%$) | |
| --- | --- | --- | --- | --- | --- | --- |
|  |  | 2.5% | 95% |  | 2.5% | 95% |
| Guayas | 0.6774 | 0.6761 | 0.6786 | 2.4253 | 2.4011 | 2.4495 |
| Manabi | 0.4794 | 0.4793 | 0.4795 | 1.7908 | 1.7844 | 1.7972 |
| El Oro | 0.6357 | 0.6351 | 0.6362 | 2.3226 | 2.3085 | 2.3367 |
| Santo Domingo | 0.5147 | 0.5142 | 0.5151 | 1.8963 | 1.8830 | 1.9097 |
| Los Rios | 0.5942 | 0.5935 | 0.5950 | 2.1718 | 2.1493 | 2.1944 |
| Sucumbios | 0.5993 | 0.5986 | 0.6000 | 2.1635 | 2.1456 | 2.1815 |
| Pichincha | 0.7221 | 0.7212 | 0.7229 | 2.6160 | 2.5920 | 2.6400 |
| Azuay | 0.5603 | 0.5600 | 0.5607 | 2.0635 | 2.0481 | 2.0789 |
| Ecuador | 0.6571 | 0.6564 | 0.6578 | 2.3977 | 2.3757 | 2.4198 |

**Table 7:** Critical values for unknown parameters in eigth provinces of Ecuador. In the case of *ρ*, and *γ*_*J*_ a value smaller than the critical value specified for the corresponding province will cause the hospitals in such province to collapse. In the case of *l*_2_, and l_3_, a value bigger than the critical value specified for the corresponding province will cause the same problem.

| Province | Critical $\rho$ | Critical $\gamma_J$ | Critical $l_2$ | Critical $l_3$ |
| --- | --- | --- | --- | --- |
| Guayas | 0.00022 | 1/4.3 | 0.45 | 0.75 |
| Manabi | 0.00072 | 1/5.0 | 0.30 | 0.50 |
| El Oro | 0.00130 | 1/3.0 | 0.30 | 0.25 |
| Santo Domingo | 0.00090 | 1/5.0 | 0.30 | 0.50 |
| Los Rios | 0.00048 | 1/9.0 | 0.48 | 0.75 |
| Sucumbios | 0.00200 | 1/4.5 | 0.29 | 0.12 |
| Pichincha | 0.00018 | 1/4.0 | 0.30 | 0.50 |
| Azuay | 0.00065 | 1/7.5 | 0.40 | 0.60 |

**Table 8:**
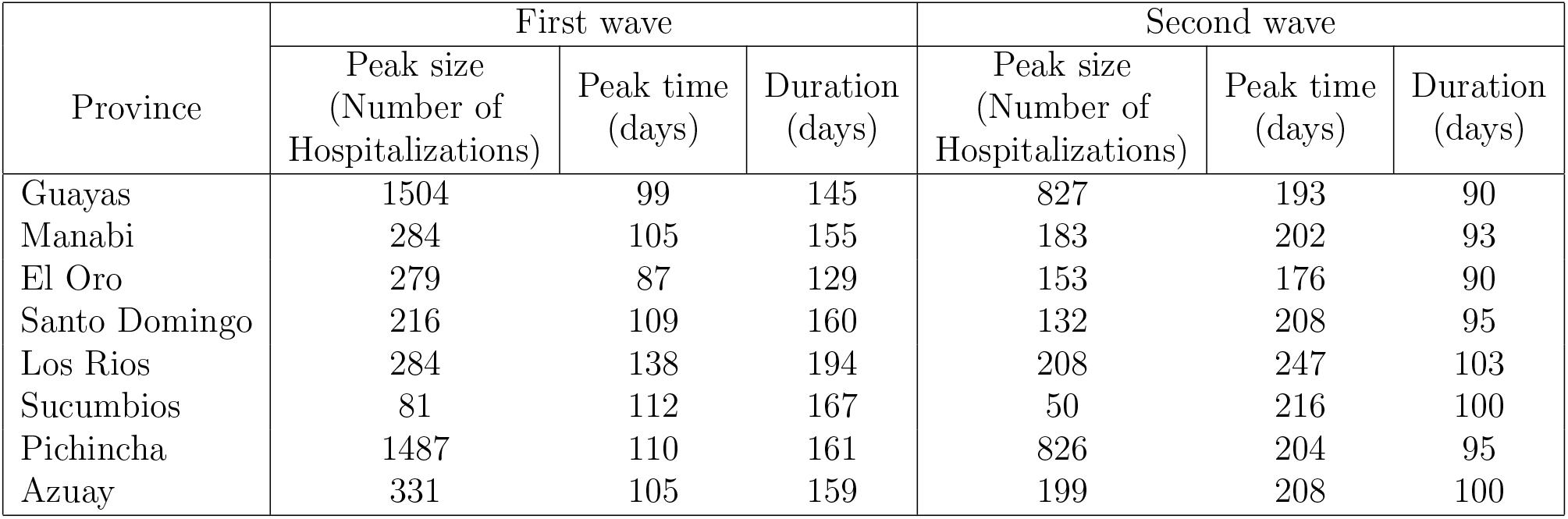
Peak size, time, and duration of the first two waves of infections in each province. The peak size accounts for the maximum number of hospitalizations happening in one day, the pike time is the number of days that have passed since the beginning of the outbreak until the peak, and the duration represents the time in days that the wave of infections lasted. The values presented in this table were obtained from the curve generated by the critical value of quarantine rate (*ρ*).

**Table 9:** Generalized growth fitting (Model 9) results for the cumulative confirmed cases of Covid-19 in Ecuador and 8 of its most affected provinces

| Province / Country | $\lambda$ | $A$ | $m$ | $p$ | RSE | $X_0$ |
| --- | --- | --- | --- | --- | --- | --- |
| Ecuador | 3.72 | 2.18 | 2.21 | 0.55 | 405.60 | 5.59 |
| Guayas | 2.62 | 3.03 | 2.33 | 0.57 | 355.80 | 13.26 |
| Manabi | 0.40 | 1.06 | 3.34 | 0.70 | 13.09 | 1.22 |
| El Oro | 0.39 | 0.65 | 3.56 | 0.72 | 9.29 | 0.21 |
| Santo Domingo | 0.26 | 1.00 | 3.27 | 0.69 | 2.95 | 1.00 |
| Los Rios | 0.93 | 1.00 | 1.97 | 0.49 | 11.52 | 1.00 |
| Sucumbios | 0.78 | 1.00 | 1.14 | 0.13 | 4.62 | 1.00 |
| Pichincha | 1.31 | 1.00 | 2.13 | 0.53 | 20.15 | 1.00 |
| Azuay | 0.71 | 1.00 | 2.10 | 0.52 | 9.45 | 1.00 |

**Table 10:** Exponential Fitting results for total confirmed deaths in Ecuador and 7 of its most affected provinces.

| Province / Country | Exponential Fitting Confirmed Deaths | | | Confidence Interval for $\lambda$ ($CI = 95\%$) | |
| --- | --- | --- | --- | --- | --- |
| | $I_0$ | $\lambda$ | $SQR$ | 2.5% | 97.5% |
| Guayas | 2 | 0.1571 | 0.9285 | 0.1419 | 0.1723 |
| Manabi | 1 | 0.2231 | 0.9226 | 0.1930 | 0.2533 |
| El Oro | 1 | 0.1370 | 0.9700 | 0.1266 | 0.1474 |
| Santo Domingo | 1 | 0.1545 | 0.9138 | 0.1244 | 0.1845 |
| Los Rios | 1 | 0.0827 | 0.8759 | 0.0693 | 0.0961 |
| Pichincha | 1 | 0.1333 | 0.9475 | 0.1200 | 0.1465 |
| Azuay | 1 | 0.0976 | 0.7182 | 0.0675 | 0.1277 |
| Ecuador | 2 | 0.2328 | 0.9422 | 0.1651 | 0.1962 |

**Table 11:** Generalized growth fitting (Model 9) results for the cumulative deaths of Covid-19 in Ecuador and 8 of its most affected provinces

| Province / Country | $\lambda$ | $A$ | $m$ | $p$ | RSE | $X_0$ |
| --- | --- | --- | --- | --- | --- | --- |
| Ecuador | 0.58 | 1.00 | 2.93 | 0.66 | 15.17 | 1.00 |
| Guayas | 0.68 | 1.00 | 2.14 | 0.53 | 14.79 | 1.00 |
| Manabi | 0.10 | 1.00 | 453.50 | 1.00 | 5.87 | 1.00 |
| El Oro | 0.11 | 1.00 | 71.24 | 0.99 | 2.39 | 1.00 |
| Santo Domingo | 0.10 | 1.00 | 2.36 | 0.58 | 2.31 | 1.00 |
| Pichincha | 0.10 | 1.00 | 730.80 | 1.00 | 3.06 | 1.00 |

**Table 12:** Number of days necessary to eliminate the epidemic in each province of Ecuador, according to Model 2.2.2, depending on the fraction of population sent into quarantine (*ρ*)

| $\rho$ | Guayas | Manabi | El Oro | Santo Domingo | Los Rios | Sucumbios | Pichincha | Azuay |
| --- | --- | --- | --- | --- | --- | --- | --- | --- |
| 1 | 44.2 | 30.6 | 33.30 | 30 | 46.2 | 30 | 39.4 | 30.6 |
| 0.1 | 63.6 | 69.5 | 72.6 | 68.8 | 58.2 | 67.6 | 62.9 | 69.5 |
| 0.01 | X | X | X | X | X | X | X | X |
| 0.001 | X | X | X | X | X | X | X | X |
| 0.0001 | X | X | X | X | X | X | X | X |
| 0.00001 | X | X | X | X | X | X | X | X |
| 0.000001 | X | 313.2 | 226.8 | 248.7 | 291.7 | 243.2 | X | 262.4 |
| 0.0000001 | 252.7 | 261.2 | 221.7 | 243.8 | 264.9 | 240.3 | 259 | 251.1 |

From the 95% confidence interval of the initial exponential growth rate (λ) presented in Table 10, 10^5^ different values of the effective transmission rate (*β*) were generated, which were then used to produce 10^5^ different values for the control reproduction number *ℛ*_*c*_. Then, 50 of these values for *ℛ*_*c*_ were used to compute the effective reproduction number (*ℛ*_*e*_) by using the formula:

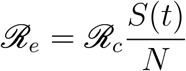

where *S*(*t*) is the number of susceptible individuals at time *t*, obtained from the numerical solution of the COVID-19 model presented in this study, and *N* is the total population size. Then, the 95% confidence interval for the mean of the 50 different values of the effective reproduction number at different time steps was computed, and the upper and lower bounds are sketched in Figure 6.

## 5 Discussion

COVID-19 disease has rapidly spread all over the world since its first identification on the 29th of December in Wuhan, China. The lives of many people have drastically changed due to the implementation of lockdown policies to restrict the spread of the virus causing the disease. Historical outbreaks have provided some clues on how quarantine and isolation type non-pharmaceutical interventions can be implemented to stop the rapid spread of the virus when treatment and vaccination are unavailable. However, unlike other diseases in past due to rapid spread nature of this disease, it has become all the more important to design robust non-pharmaceutical interventions for resource limited countries. Ecuador has implemented quarantine and isolation measures since March 17th, 2020 but with low to moderate efficacy. In this study, three different types of models were presented as possible tools to analyze and predict the spread of COVID-19 in Ecuador and its provinces.

The first model, exponential epidemic growth model, was fitted to the cumulative confirmed and mortality data for eight provinces of Ecuador and the whole country, obtaining the initial exponential growth rate along with its corresponding 95% confidence interval. The fitting of the confirmed cases produced an adjusted R squared (SQR) around 0.89 whereas that of the cumulative deaths produced a SQR around 0.94, meaning that a great part of the observed variation in the confirmed cases and deaths can be explained by the time that has passed since the first cases. However, the fitting done to the cumulative confirmed cases can be limited because the number of COVID-19 cases in Ecuador may be underreported. The reported cumulative deaths are closer to reality since the majority of COVID-19 related deaths occur in hospitals, with some exceptions.

The second model, generalized epidemic growth model, was also fitted to the cumulative confirmed cases and deaths, obtaining the growth rate corresponding to eight Ecuador provinces and the whole country. The main results obtained from this model were that the growth rate in the cumulative confirmed cases and deaths was quadratic in most of the provinces, with the exception of Manabi, El Oro, and Pichincha that had exponential growth rate in the number of deaths.

The third model, presented in section 2.2.2, is a compartmental model that incorporates quarantine and isolation as interventions. From the simulations made with this model, we found that the implementation of quarantine measures causes the epidemic to evolve slower, since many people are taken out of the susceptible compartment and sent into quarantine, and then slowly come back to the susceptible compartment. This slower evolution in the dynamics of the disease, caused by the implementation of quarantine, is positive because it avoids overloading the hospitals as seen in Figure 2a. This Figure shows that the quarantine rate per hospitalized case (*ρ*) is under 0.0002, which means that if 2 of each 10000 susceptible individuals go to quarantine per day, it can cause the hospital to collapse in about 120 days in Guayas. The number of deaths also increases drastically depending on this quarantine rate as depicted in Figure 2b. The symptomatic infectious quarantine rate (*γ*_*J*_) has a similar effect on the number of hospitalizations (Figure 3a), and cumulative deaths (Figure 3b) as *ρ*, which are increased when a smaller value for *γ*_*J*_ is used. In other words, the longer the symptomatic infectious individuals stay in the *I* compartment, the more the number of new infections increases, and therefore the hospitalizations and deaths also increase. The proportion of symptomatic infectious people that are not strictly isolated (*l*_2_) has also a strong impact on the number of hospitalizations (Figure 4a) and cumulative deaths (Figure 4b) since the more the symptomatic infectious people leak from isolation the more people they infect. Finally, the proportion of people that are not strictly hospitalized due to limited equipment (*l*_3_) has a similar effect as *l*_2_ on the dynamics of the disease, since the more the people in need of hospitalization do not get the attention they need the more people they infect. In summary, there is a tradeoff between high quarantine (and isolation) rate and leakage in the interventions. *High intervention rates is not always beneficial, unless we can fine tune (that is, contained) the leakage of infection through these measures*.

**Figure 3:**
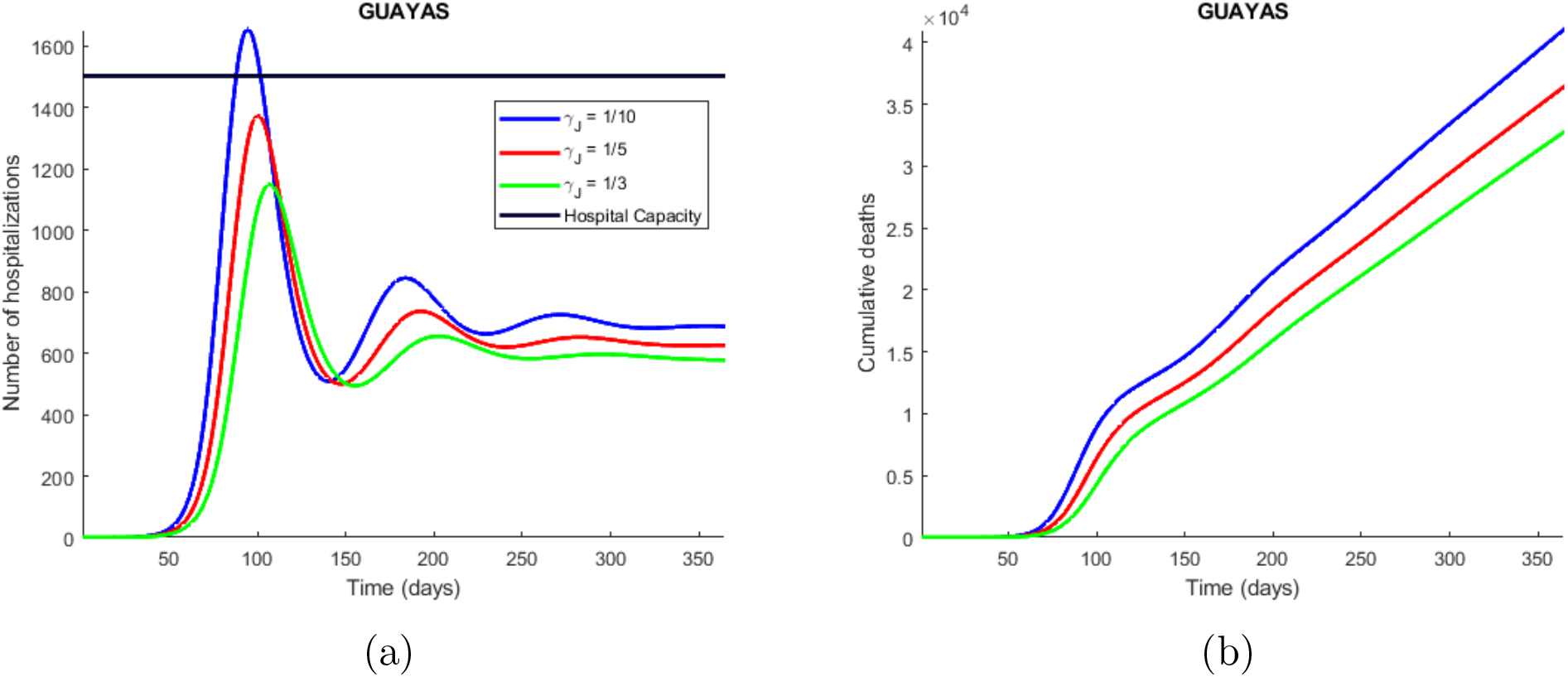
Time series plot of the number of hospitalizations (a), and cumulative deaths (b) predicted by the model. The simulations were done for Guayas province with the parameters values presented in Table 2 and varying the symptomatic infectious quarantine rate (*γ*_*J*_) between 1/3 (people showing COVID-19 symptoms are sent into isolation within 3 days) and 1/10. The legend specifies the values for the quarantine rates that produce the corresponding curves, considering the explained meaning of such values. The horizontal line represents the hospitals capacity in the province.

**Figure 4:**
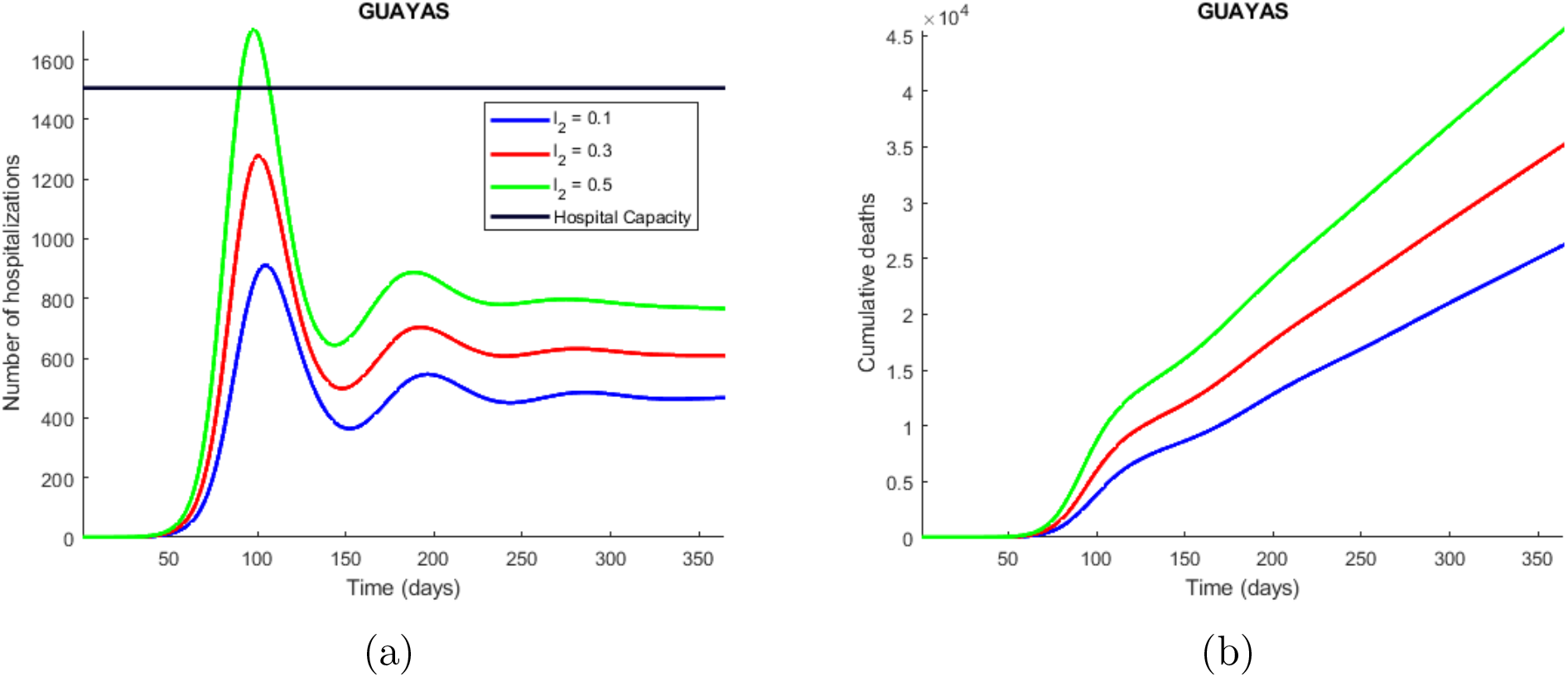
Time series plot of the number of hospitalizations (a), and cumulative deaths (b) predicted by the model. The simulations were done for Guayas province with the parameters values presented in Table 2 and varying the proportion of symptomatic infectious people that are not strictly isolated (*l*_2_) between 0.1 (10 % of the symptomatic infectious people are not strictly isolated) and 0.5. The legend specifies the values for the studied parameter that produce the corresponding curves, considering the explained meaning of such values. The horizontal line represents the hospitals capacity in the province.

**Figure 5:**
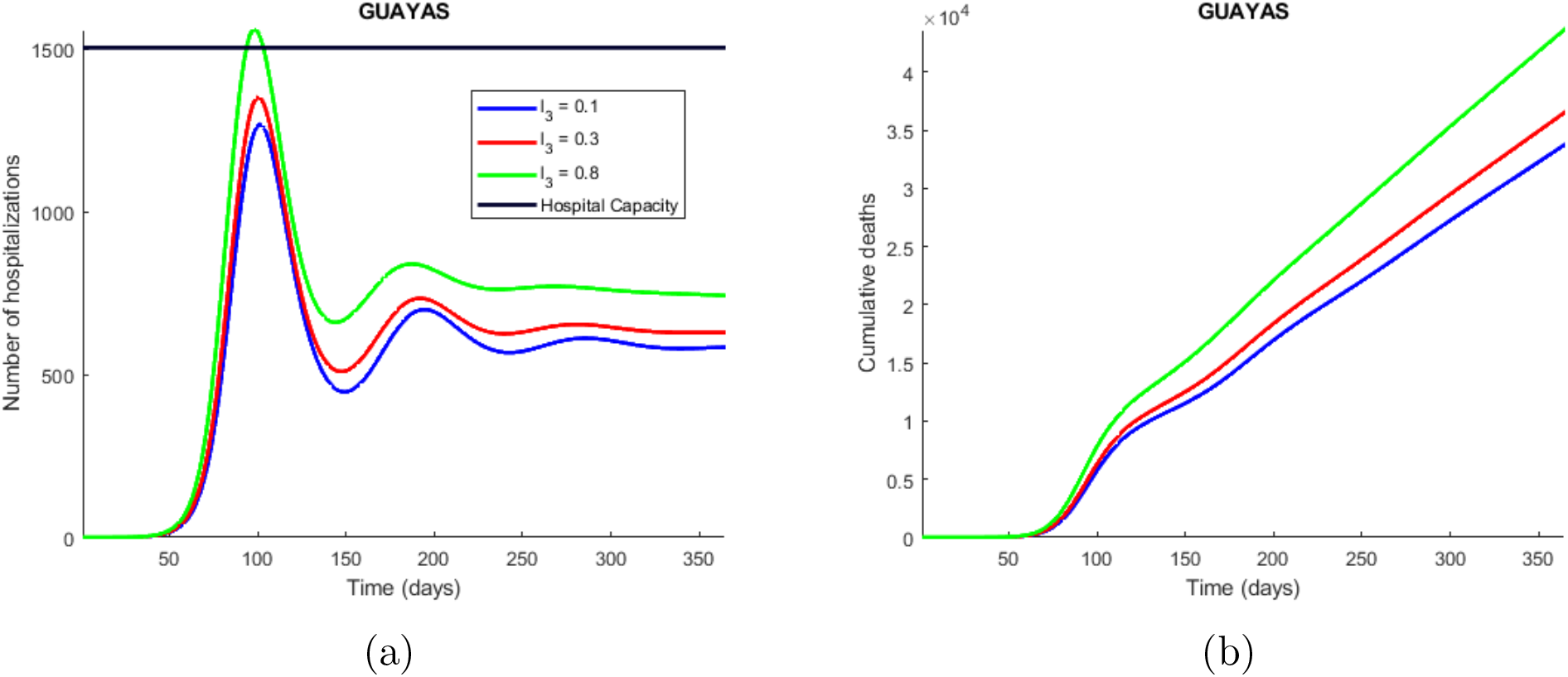
Time series plot of the number of hospitalizations (a), and cumulative deaths (b) predicted by the model. The simulations were done for Guayas province with the parameters values presented in Table 2 and varying the proportion of people that are not strictly hospitalized due to limited equipment (*l*_2_) between 0.1 (10 % of the symptomatic infectious people are not strictly isolated) and 0.5. The legend specifies the values for the studied parameter that produce the corresponding curves, considering the explained meaning of such values. The horizontal line represents the hospitals capacity in the province.

**Figure 6:**
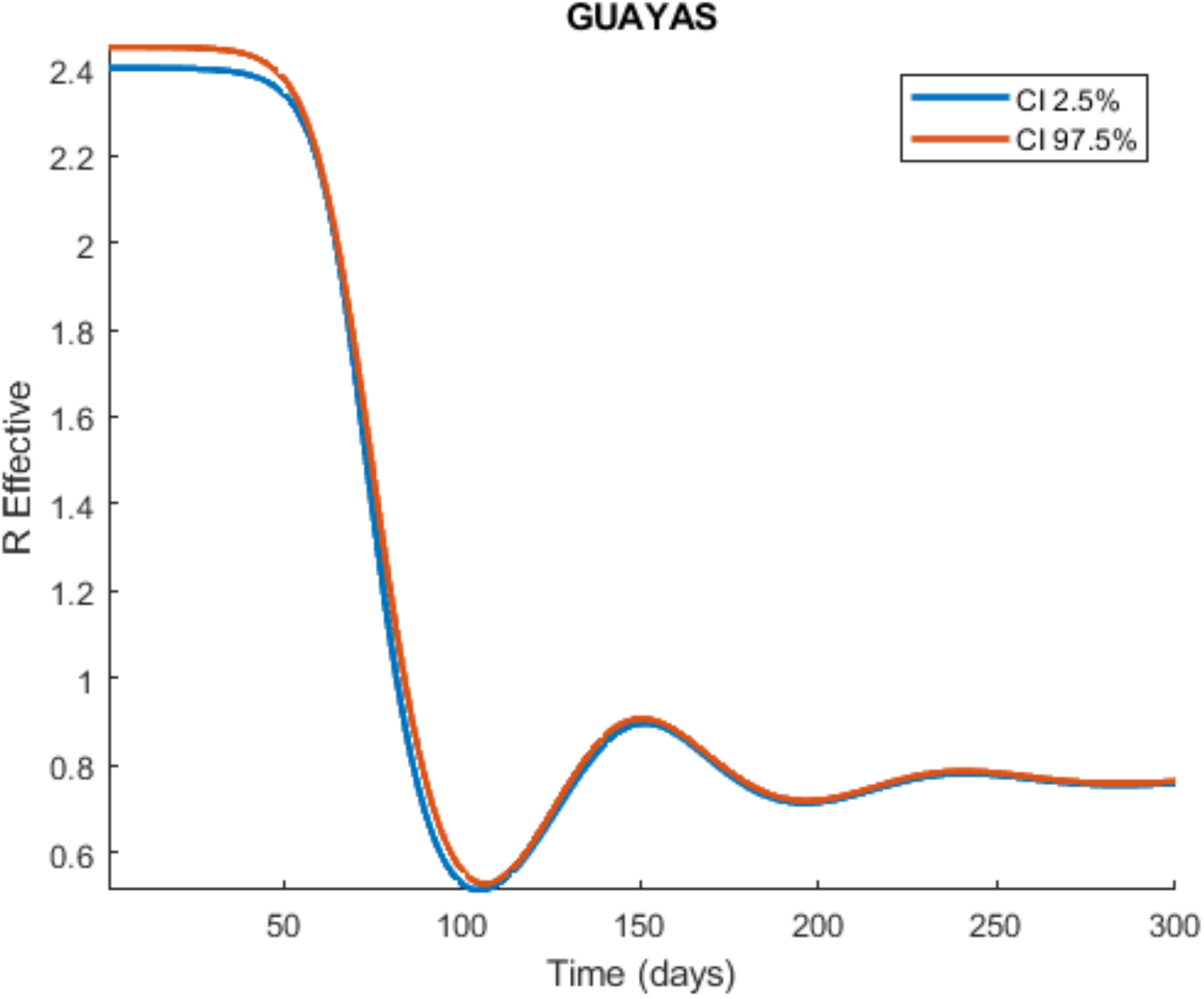
Time series plot of the effective reproduction number (*ℛ*_*e*_) for the Guayas province. The curves represent the upper and lower bounds of the 95% confidence interval of *ℛ*_*e*_. These simulations were made from the COVID-19 model presented in this study, using the parameter values from Table 2 and sampling the four unknown parameters.

**Figure 7:**
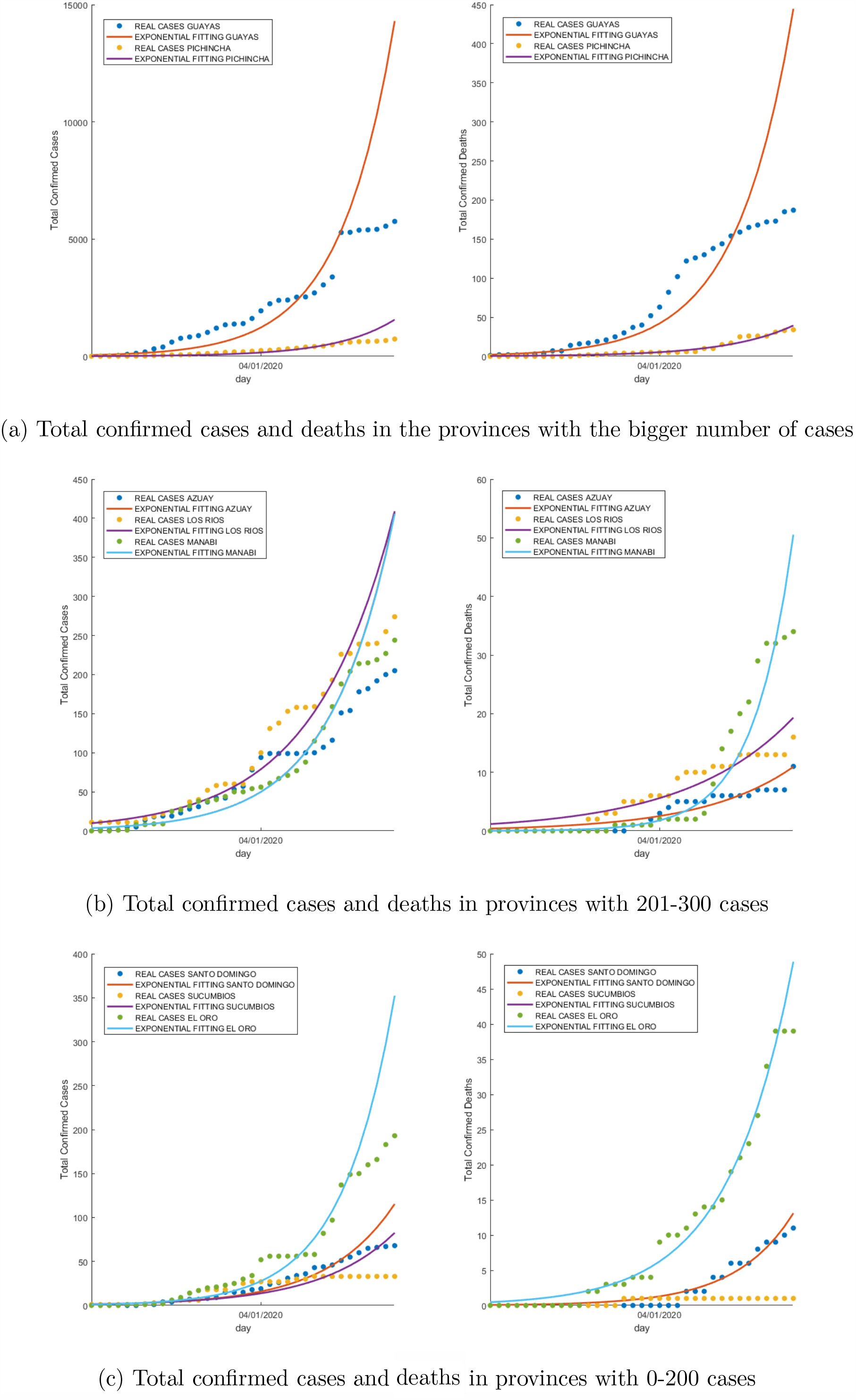
Total confirmed cases and deaths in the 8 most affected provinces

**Figure 8:**
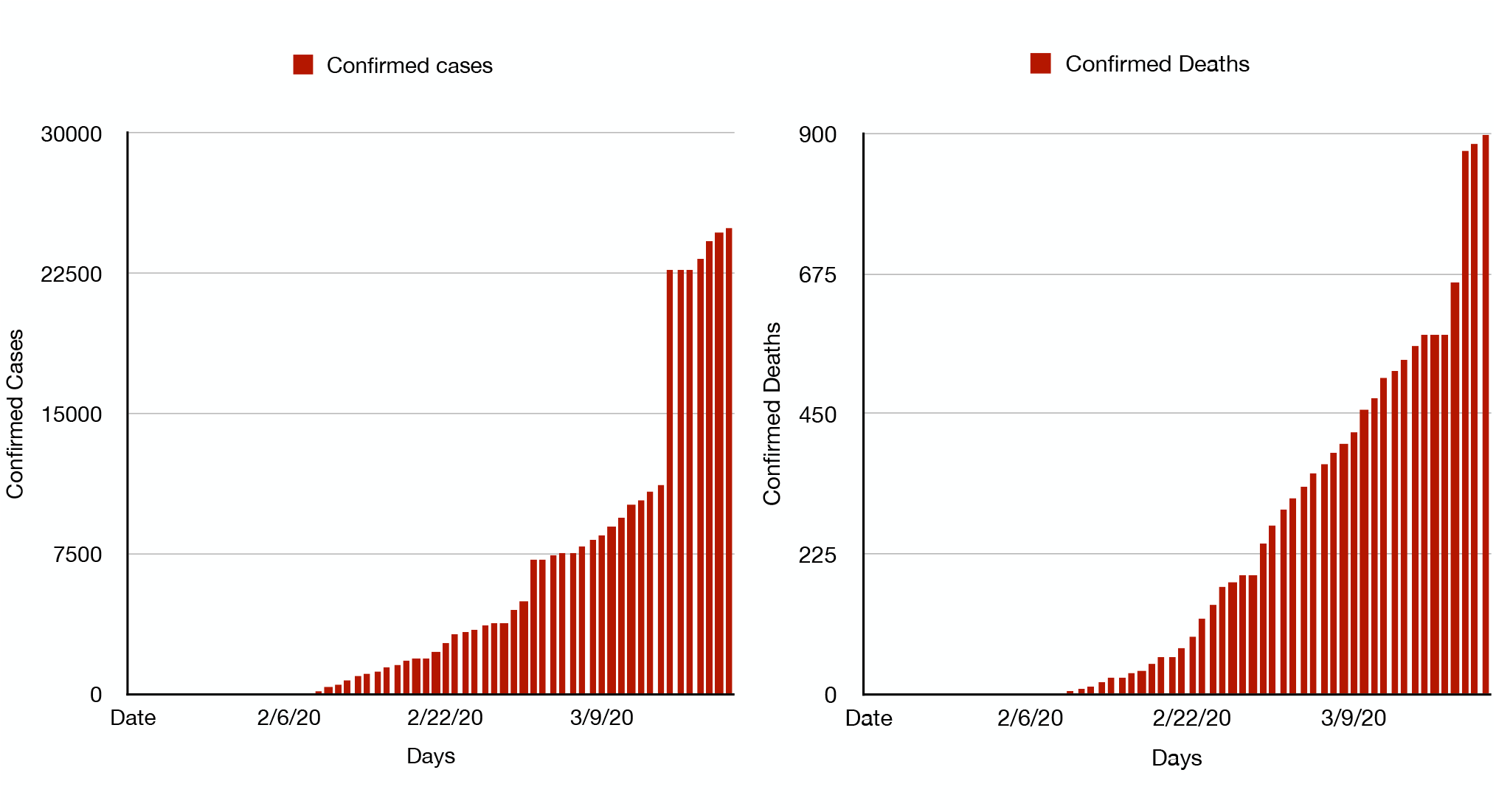
Evolution of confirmed cases and deaths in Ecuador

**Figure 9:**
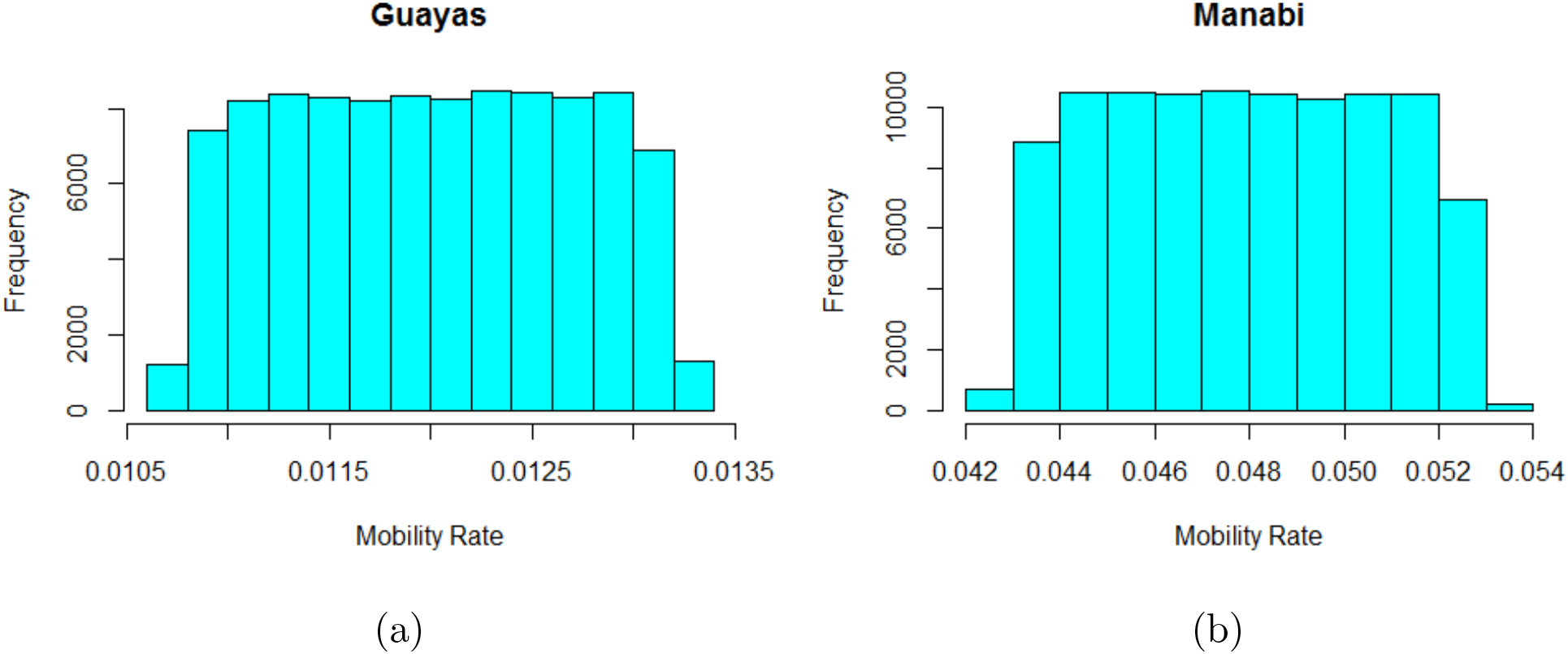
Histogram plot for the mobility rate of Guayas (a) and Manabi (b) provinces, generated from the equation ?? by varying the number of inter-provincial buses available in each province and the number of people such buses transport in a day.

**Figure 10:**
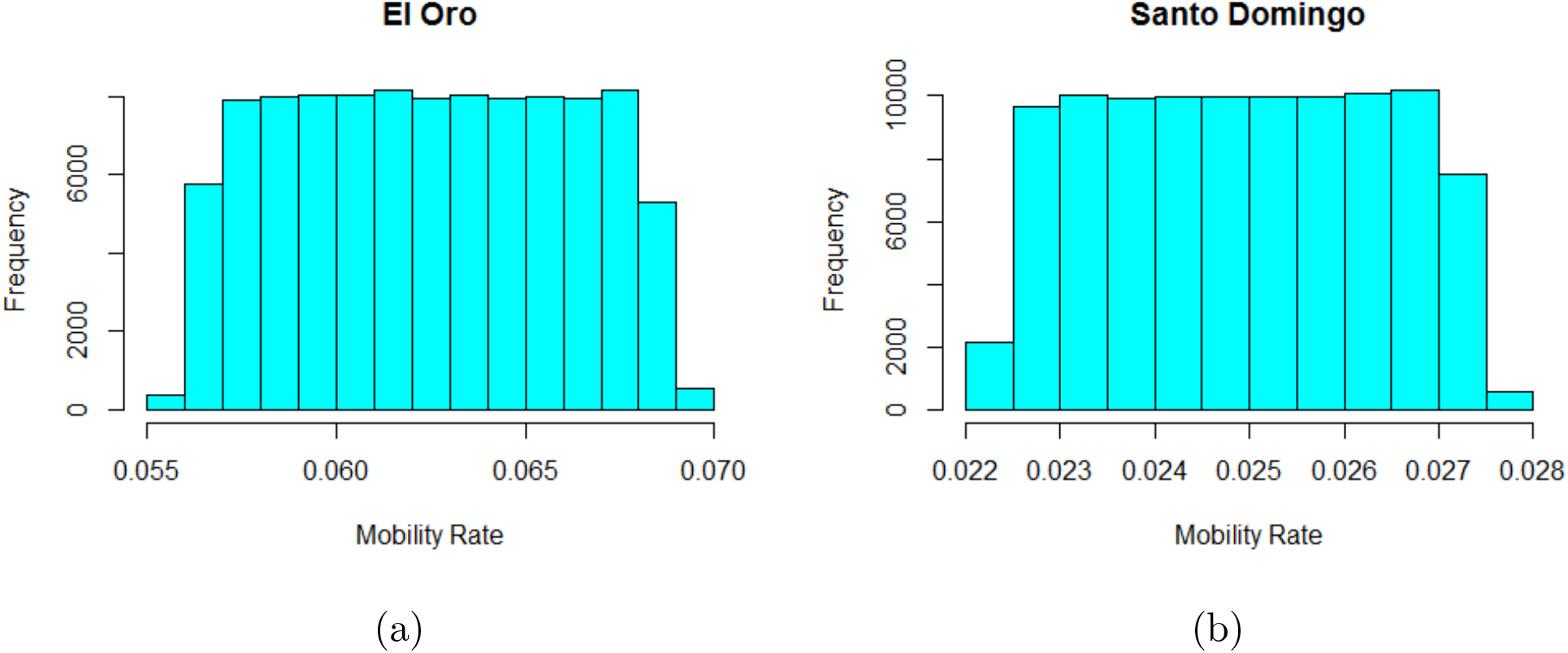
Histogram plot for the mobility rate of El Oro (a) and Santo Domingo (b) provinces, generated from the equation ?? by varying the number of inter-provincial buses available in each province and the number of people such buses transport in a day.

**Figure 11:**
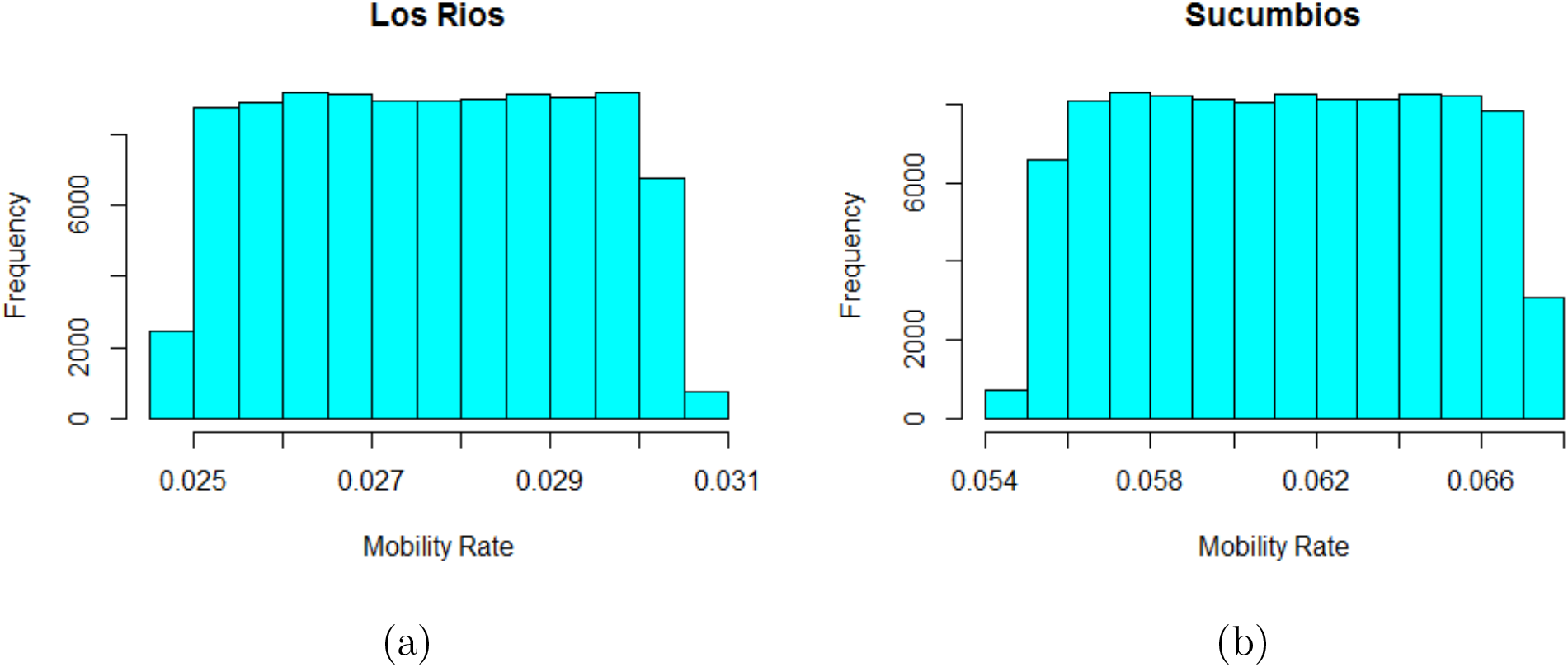
Histogram plot for the mobility rate of Lor Rios (a) and Sucumbios (b) provinces, generated from the equation ?? by varying the number of inter-provincial buses available in each province and the number of people such buses transport in a day.

**Figure 12:**
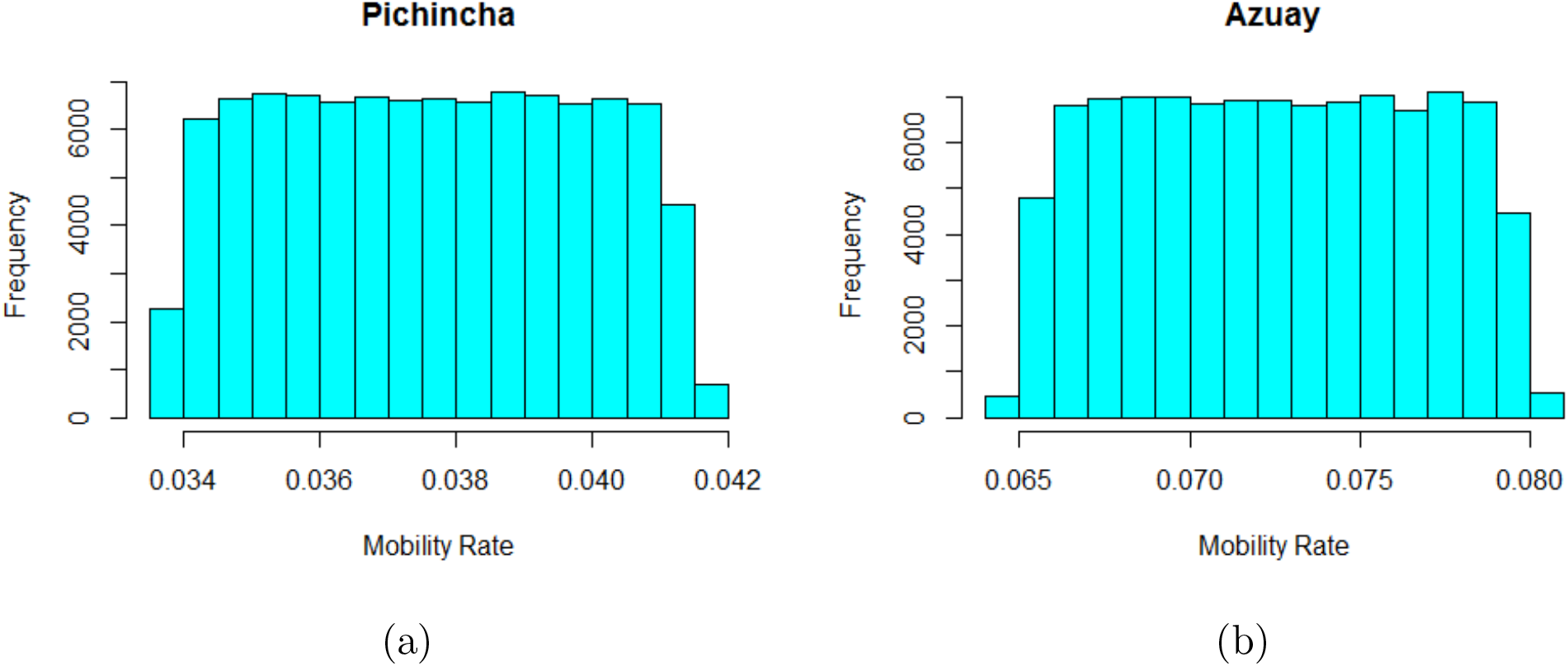
Histogram plot for the mobility rate of Pichincha (a) and Azuay (b) provinces, generated from the equation ?? by varying the number of inter-provincial buses available in each province and the number of people such buses transport in a day.

**Figure 13:**
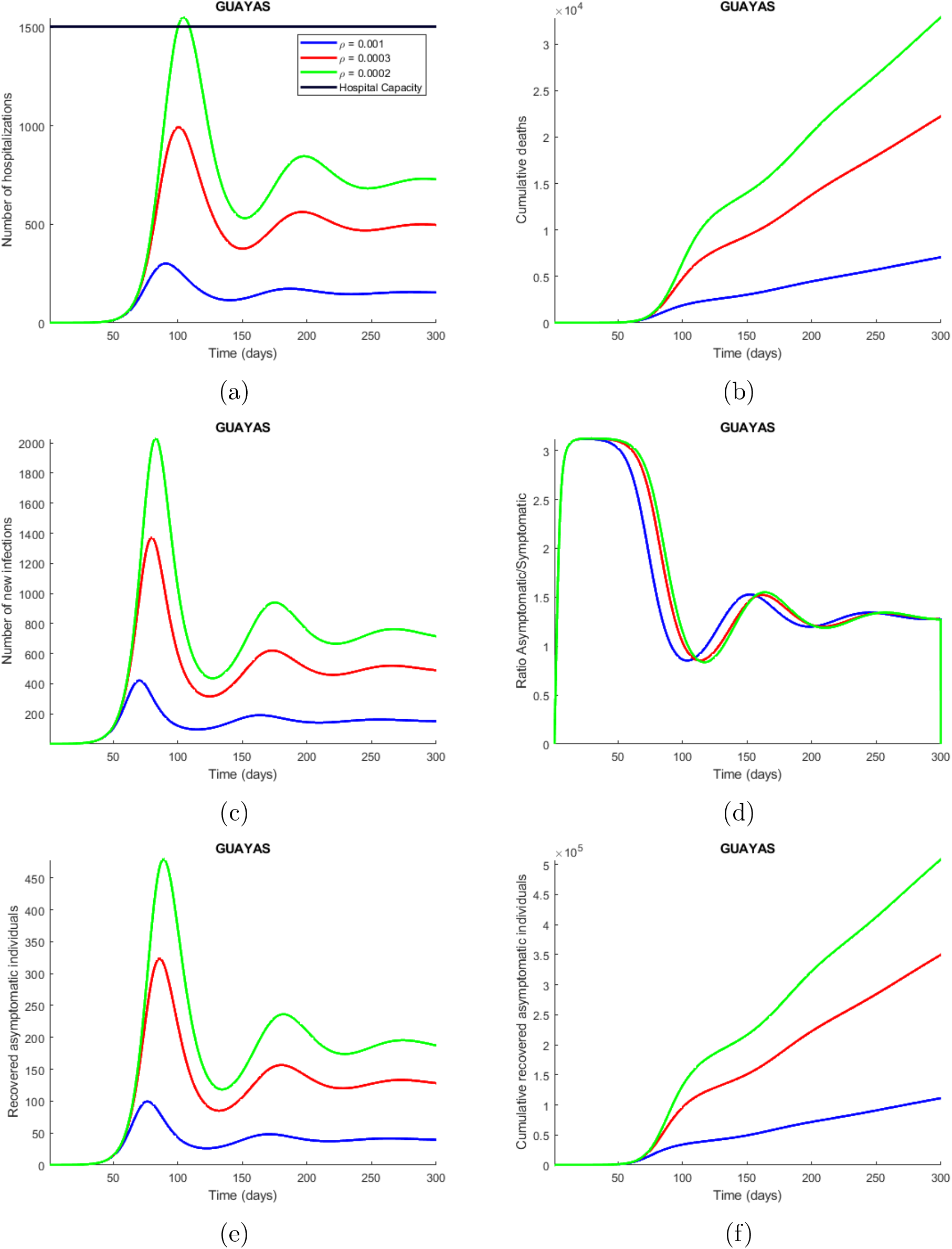
Time series plot of the number of hospitalizations (a), cumulative deaths (b), number of new infections (c), ratio asymptomatic/symptomatic (d), number of new recovered asymptomatic individuals (e) and cumulative recovered asymptomatic individuals (f) predicted by the model. These simulations were done for Guayas province with the parameters values presented in Table 2 and varying the quarantine rate (*ρ*) between 0.001 (10 of each 10000 influenced susceptible individuals go to quarantine) and 0. 0002. The legend specifies the values for the quarantine rates that produce the corresponding curves, considering the explained meaning of such values. The horizontal line represents the hospitals capacity in the province.

**Figure 14:**
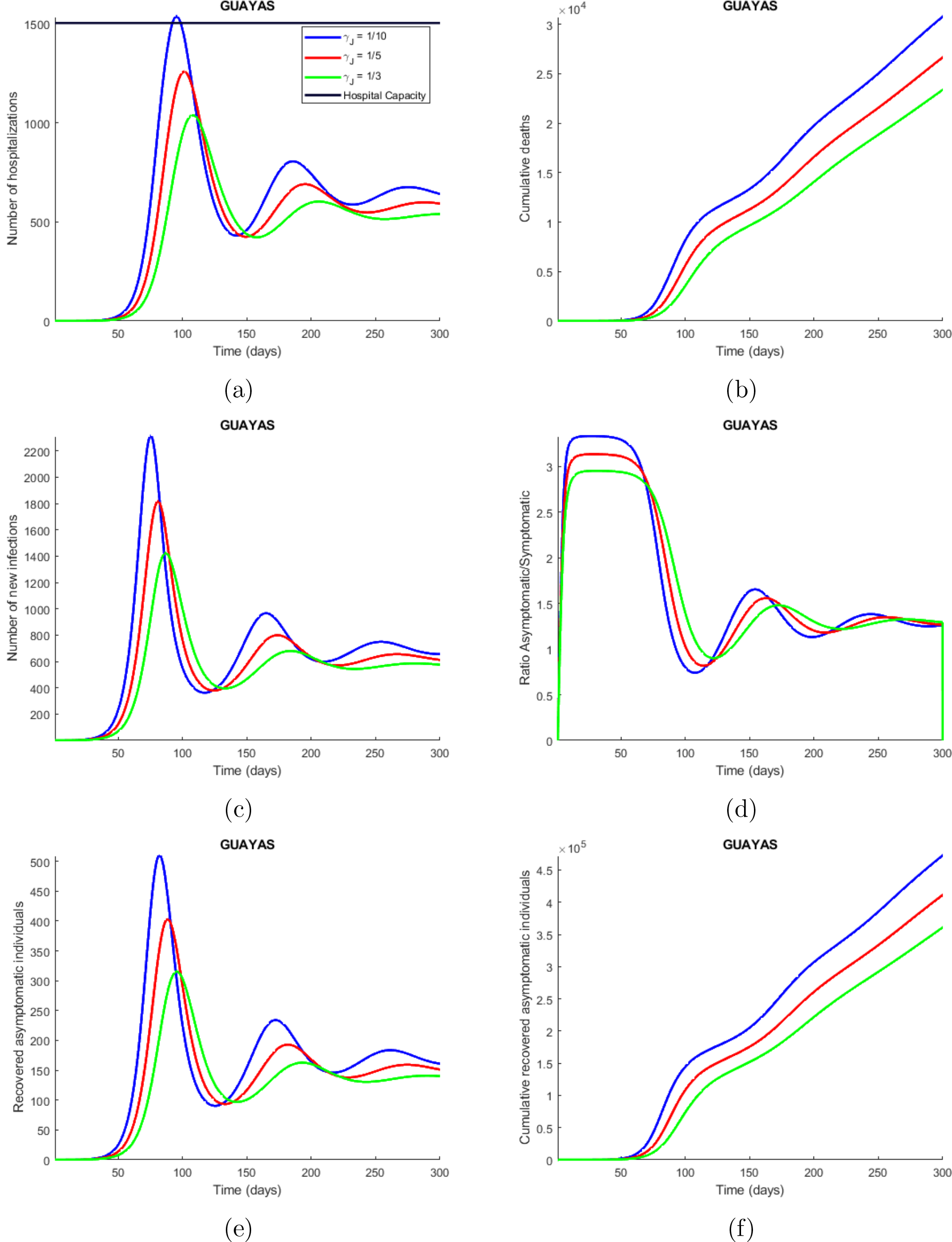
Time series plot of the number of hospitalizations (a), cumulative deaths (b), number of new infections (c), ratio asymptomatic/symptomatic (d), number of new recovered asymptomatic individuals (e) and cumulative recovered asymptomatic individuals (f) predicted by the model. These simulations were done for Guayas province with the parameters values presented in Table 2 and varying the symptomatic infectious quarantine rate (*γ*_*J*_) between 1/3 (people showing COVID-19 symptoms are sent into isolation within 3 days) and 1/10. The legend specifies the values for the quarantine rates that produce the corresponding curves, considering the explained meaning of such values. The horizontal line represents the hospitals capacity in the

**Figure 15:**
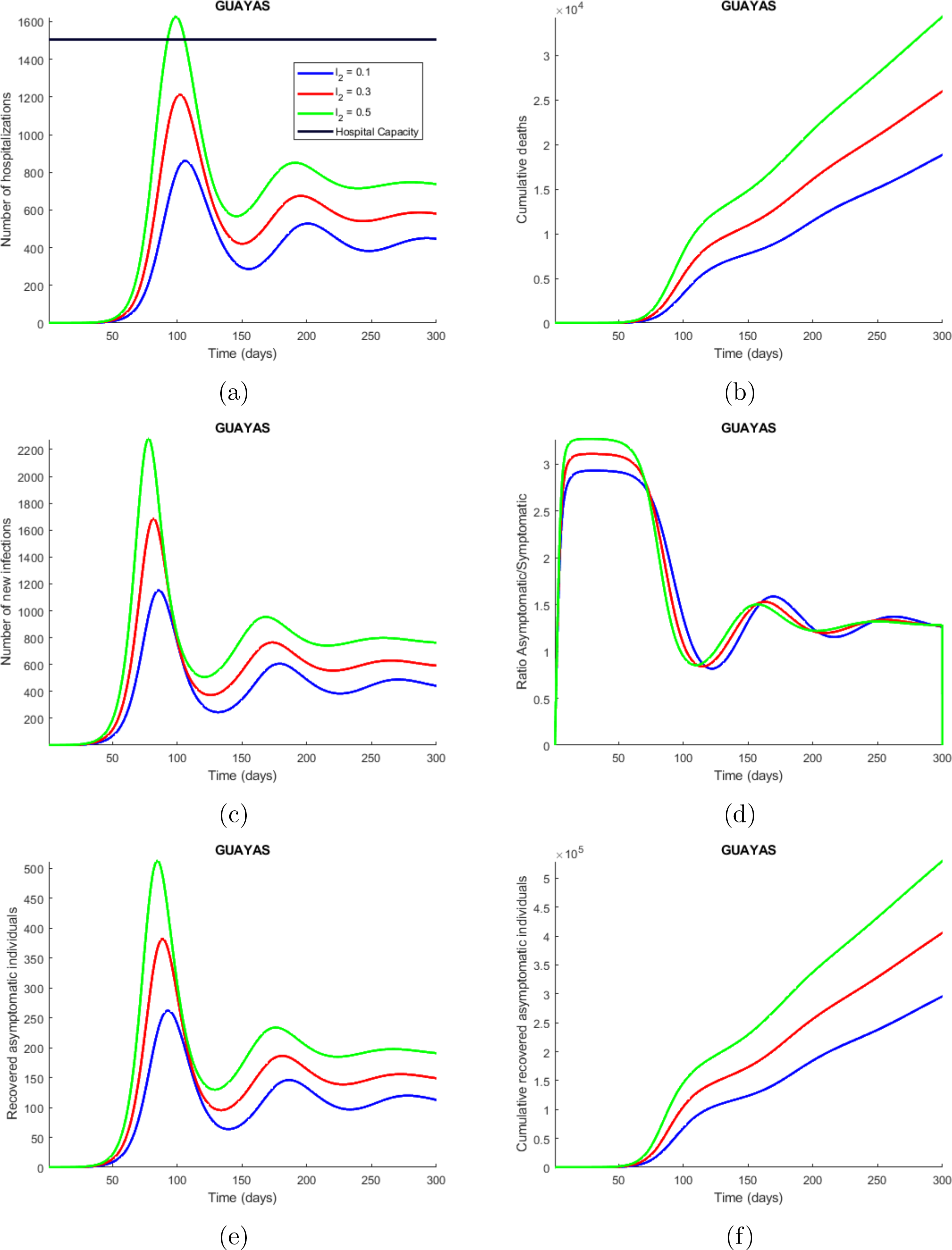
Time series plot of the number of hospitalizations (a), cumulative deaths (b), number of new infections (c), ratio asymptomatic/symptomatic (d), number of new recovered asymptomatic individuals (e) and cumulative recovered asymptomatic individuals (f) predicted by the model. These simulations were done for Guayas province with the parameters values presented in Table 2 and varying the proportion of symptomatic infectious people that are not strictly isolated (*l*_2_) between 0.1 (10 % of the symptomatic infectious people are not strictly isolated) and 0.5. The legend specifies the values for the studied parameter that produce the corresponding curves, considering the explained meaning of such values. The horizontal line represents the hospitals capacity in the province.

**Figure 16:**
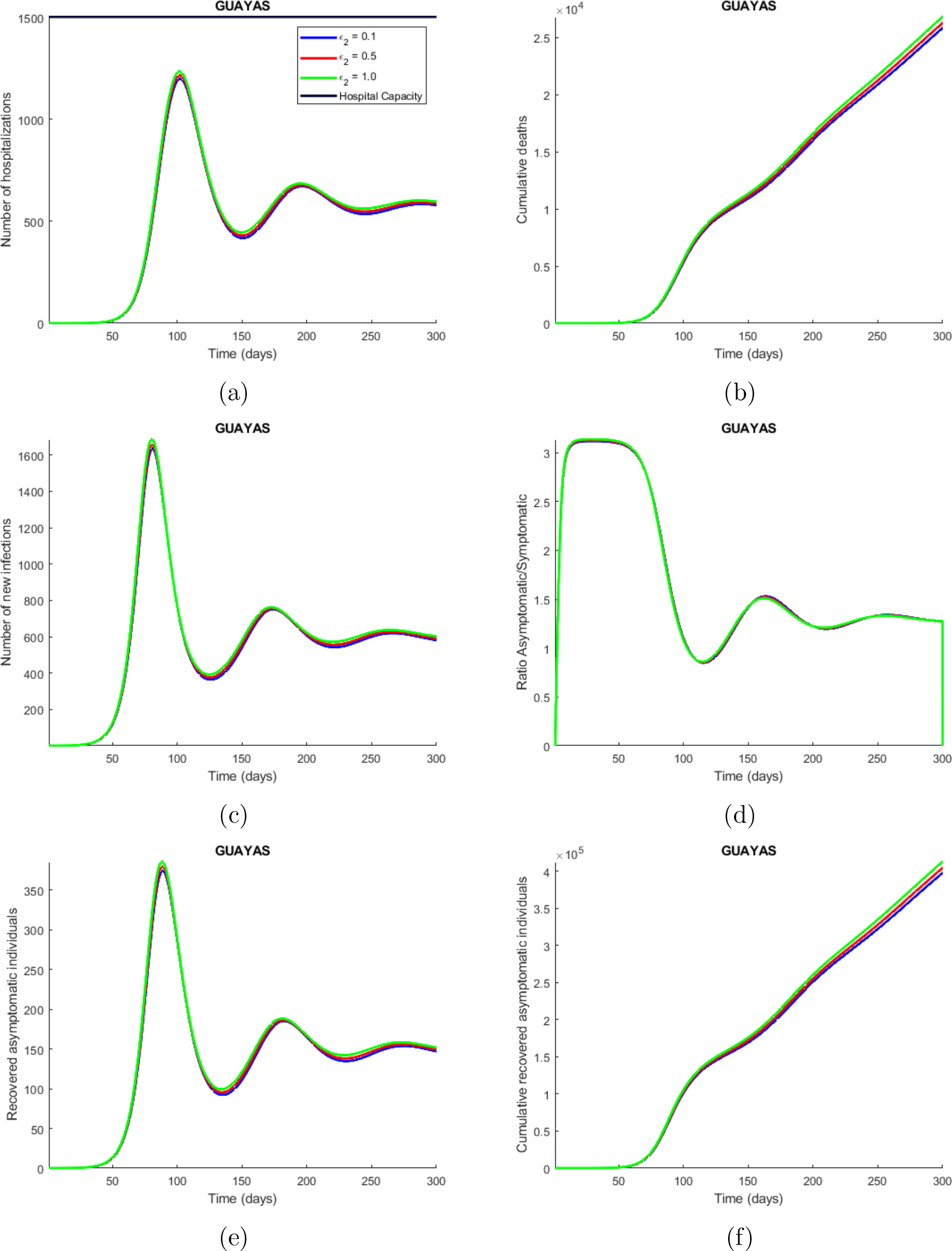
Time series plot of the number of hospitalizations (a), cumulative deaths (b), number of new infections (c), ratio asymptomatic/symptomatic (d), number of new recovered asymptomatic individuals (e) and cumulative recovered asymptomatic individuals (f) predicted by the model. These simulations were done for Guayas province with the parameters values presented in Table 2 and varying the infectiousness of hospitalized individuals (∈_2_) between 0.1 (hospitalized individuals are only 10 % infectious as compared with those symptomatic individuals that have not been detected) and 1. The legend specifies the values for the studied parameter that produce the corresponding curves, considering the explained meaning of such values. The horizontal line represents the hospitals capacity in the province.

The effective reproduction number for the Guayas province is around 2.4 at the beginning of the epidemic, and drastically decreases in the first 100 days, to increase again and stabilize around 0.8 in 300 days after the beginning of the epidemic (see Figure 6). These results are being caused by the quarantine, because at the beginning of the epidemic, the susceptible population is rapidly drained by the other compartments, specially *Q*_1_, which is the quarantine for the susceptible people. The effective reproduction number in all the provinces are significantly dropping, indicating the end of the firs outbreak. The damped oscillations observed in the different figures, including the one presented for the effective reproduction number are caused by the quarantine rate because it drains the susceptible population very fast at the beginning, but then it starts to slowly come back, causing new outbreak after every certain amount of time. In the case of no control interventions, the final epidemic size will be around one million individuals (see C.3).

The implementation of the proposed models critically depends of the available data, mainly the number of confirmed cases, the number of deaths, and information about health care system resources related to this respiratory disease (ventilators, beds, etc) at provincial and national level. To the best of our knowledge, this study captures different scenarios of the COVID-19 outbreak in Ecuador. Here, we provide mathematical tool for the decision-makers to effectively implement non-pharmaceutical interventions. Indeed, the implementation of quarantine, in an appropriate proportion, can flatten the curve of new infections and avoid the collapsing of the health care system. But as the cases will reduce, the social distancing measure may also get relaxed and thus creating a potential for second wave of the outbreak.

## Data Availability

All data are included in the manuscript

## Acknowledgements

The last author acknowledge the partial support from NSF grant 1918614.

## Authors Contribution

A.M. conceived of the presented idea. C.B., J.C. and A.M. developed the theory, carried out analytical methods and performed the computations. C.M., J.S., E.M. and A.M. verified the computational scenarios and methods as well as contributed to the interpretation of the results. C.B., J.C., and C.M. took the lead in initial writing the manuscript. A.M. was in charge of overall direction and planning and supervised its findings. All authors discussed the results and contributed to the final manuscript.

## A Appendix

## B Appendixs

### B.0.1 Exponential Models

#### Exponential Epidemic Growth Model

The exponential growth rate of an epidemic is a key measure of the severity at which the epidemic can impact a population, and it is closely related to the basic reproduction number (*R*_0_) [13]. At the beginning of an epidemic, the number of cases tend to grow exponentially, especially if no strict measures are taken. So, this model intends to explain the growth in the number of cases at the beginning of an outbreak. The model that describes an exponential growth rate in the number of confirmed cases of infected people is:

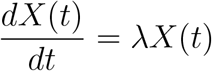

which has the solution:

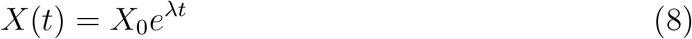

where *X*(*t*) is the number of cases at time *t, X*_0_ is the number of initial cases, and, λ is the growth rate. *ln* (*X*(*t*)) and t have a linear relationship during the initial growth phase, hence:

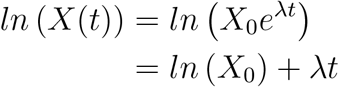

This linear relationship allows us to fit a linear model to the natural logarithm of the data about confirmed cases and obtain the initial exponential growth rate, λ and the initial number of cases *X*_0_.

#### Generalized Epidemic Growth Model

The initial cumulative growth pattern of outbreaks has been widely studied using models that principally assume exponential growth dynamics when no countermeasures are taken [15]. However, the cumulative number of cases, *X*(*t*), often has been observed to grow slower than exponential, with infectious diseases that spread via close contacts, in scenarios that include restricted contact among individuals [14]. Hence, we consider a simple generalized model that relaxes the exponential growth rate as assumption:

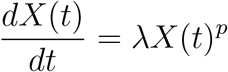

where, *X*(*t*) is the cumulative number of cases at time t,, λ is a positive parameter denoting the growth rate (1/time), and *p* ∈ [0, 1] is a deceleration of growth parameter. If *p* = 0, this equation describes constant incidence over time and the cumulative number of cases grows linearly while *p* = 1 models exponential growth dynamics (i.e., Malthus equation). Intermediate values of p between 0 and 1 describe sub-exponential (e.g., polynomial) growth patterns. For example, if *p* = 1/2 incidence grows linearly while the cumulative number of cases follows a quadratic polynomial. If *p* = 2/3 incidence grows quadratically while the cumulative number of cases fits a cubic polynomial [15]. For sub-exponential growth (i.e., 0 < *p* < 1) the solution of this equation is given by the following polynomial of degree m:

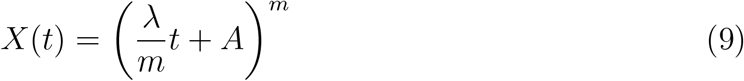

where, *m* is a positive integer, the deceleration of growth parameter is given by *p* = 1 − 1/*m* and 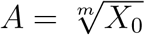

#### Power-law with exponential cut-off

Power-law functions are impressively ubiquitous, but they are not the only form of broad distribution that have been observed for a population growth, including for initial growth of an epidemic. For example, it could be that the function starts out as an exponential and ends up as a power law or vice versa. In other words, the distribution of cases may follow a exponential only for part of the time-axis range but are cut off at high values of time. That is, above some value they deviate from the exponential and rises slowly towards the peak of an outbreak. This type of functions can be used to estimate the inflection points of a function, which could represent the time when interventions starts to impact the outbreak growth. Such model (similar to ones used in ecology [22]) can be represented as

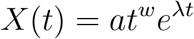

or

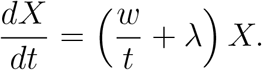

where, for example, *t* is the time since the beggining of an outbreak, *X* is the cumulative incidence, and *a*, λ, and *w* are parameters to be estimated from the observed data. Using simple computations, it can be obtained that the inflection point of the function occurs at *t* = −*w*/λ, where *w* > 0, but, λ < 0.

### B.1 Summary of Exponential Models

The general form of all the exponential models can be written using following function

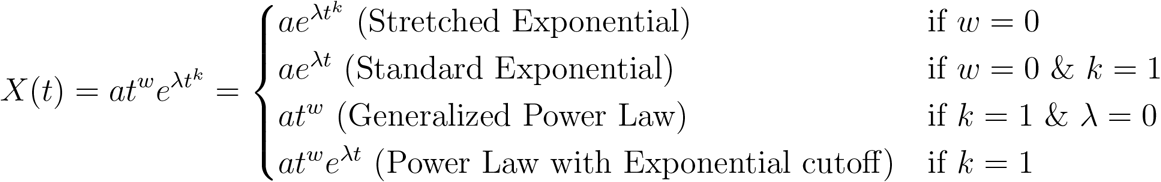

Typically, the outbreak is believed to follow growth in three parts until it reaches its peak: (*i*) initial polynomial growth (*ii*) middle exponential growth and (*iii*) eventual linear growth. That is,

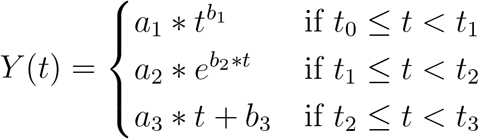

### B.2 Full Model

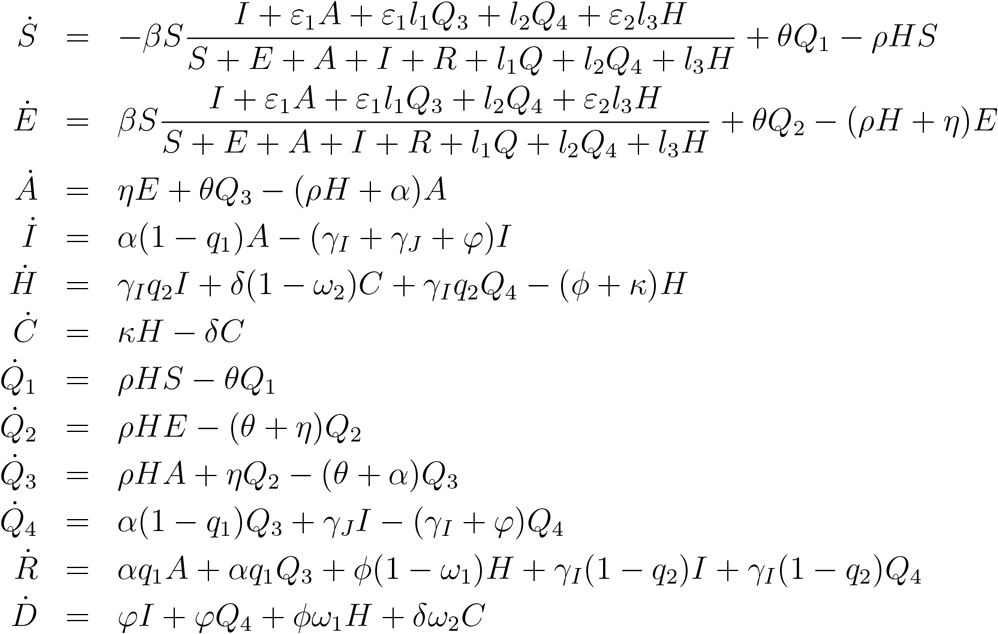

## C. Appendix

### C.1 Basic Reproduction Number for the Baseline Model (SEAIR)

The basic reproduction number for the baseline and for the complete models is computed by using the next generation operator method [21]. The matrix ℱ of the new infection terms 𝒱 and of the remaining transfer terms associated to the baseline model are given by

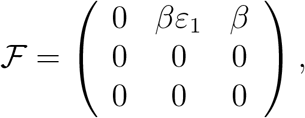

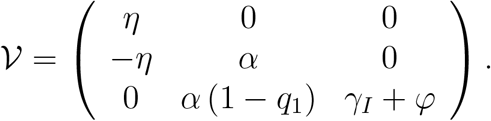

Then, the basic reproduction number, named *ℛ*_0_, for the baseline model is given by

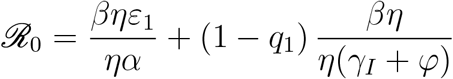

### C.2 Basic Reproduction Number for the Full Model

In an analogous way as the computation of the basic reproduction number for the complete model we will have

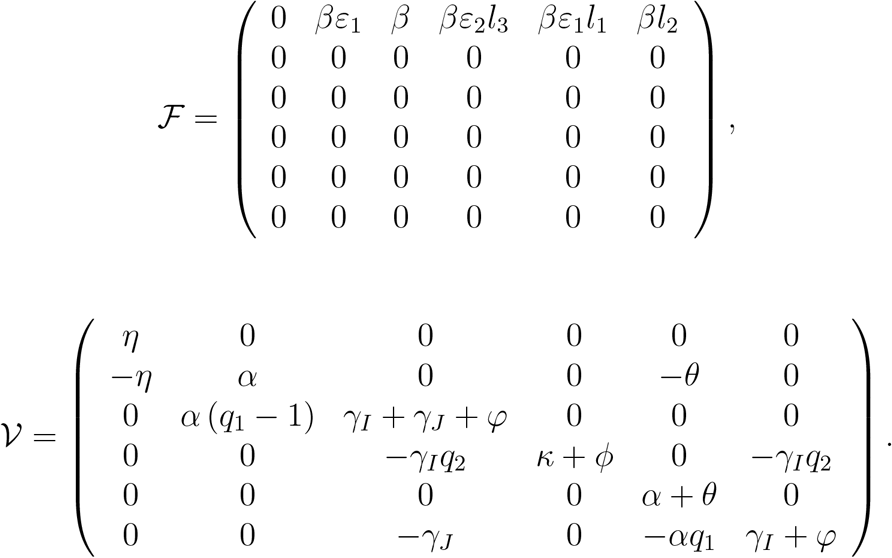

The control reproduction number, denoted as *ℛ*_*c*_, is given by

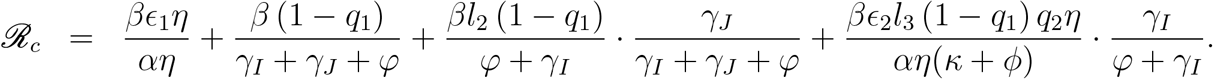

### C.3 Final Epidemic Size Baseline Model

The force of infection for the baseline model is

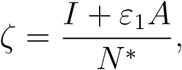

where *N*^*^ is given by

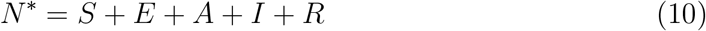

Let’s assume that the general population decreases by death induced by the disease in a proportion 0 <*p* ≪ 1 through the outbreak, i.e. *N*^*^ = (1 − *p*)*S*_0_, for COVID-19 *p* ∈ (0.04, 0.15), [1, 2]. Thus,

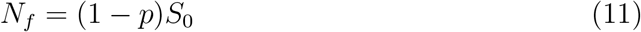

Proceeding as Brauer and Castillo-Chávez in [23], we are going to estimate the final epidemic size in Ecuador. Let’s assume,

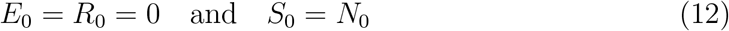

Then, we would have *S*′(*t*) + *E*′(*t*) = (*S*(*t*) + *E*(*t*)) ′ = −η*E*(*t*) ≤ 0. The function *S*(*t*) + *E*(*t*) is a smooth decreasing function and therefore tends to a limit as *t* → ∞. Also, the derivative of a positive decreasing function tends to zero, therefore η*E* → 0. Since, η > 0 then *E* → 0, i.e. E_∞_ = 0. Thus,

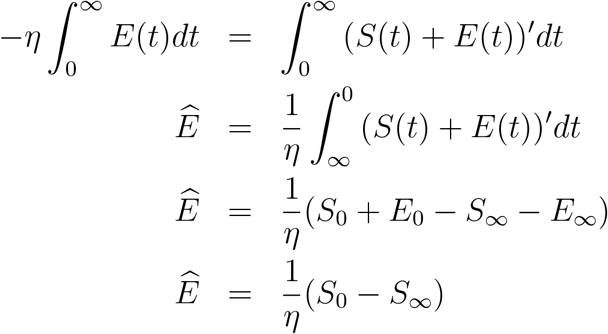

We also would have *S*′(*t*) + *E*′(*t*) + + *A*′ (*t*) = (*S*(*t*) + *E*(*t*) + *A*(*t*)) ′ = − α*A* ≤ 0. The function *S* + *E* + *A* is a smooth decreasing function and therefore tends to a limit as t → 0. Also, the derivative of a positive decreasing function tends to zero, therefore for α > 0 we have that A → 0. Thus,

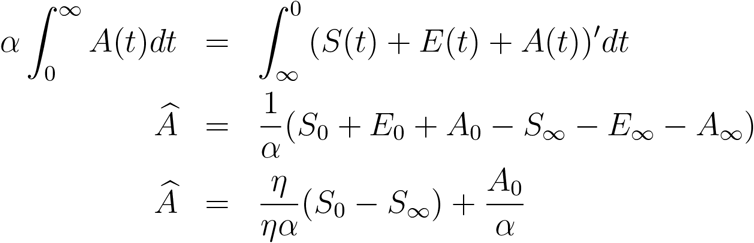

In the same way, we have *S*′(*t*) + *E*′(*t*) + + *A*′ (*t*) + *I* ′ (*t*) = (*S*(*t*)+ *E*(*t*)+ *A*(*t*)+ *I*(t)) ′ = α(1 − *q*_1_)*A*− (*γ* + *φ*)*I* ≤ 0. The function S + E + A+ I is a smooth decreasing function and therefore tends to a limit as t → ∞. Also, the derivative of a positive decreasing function tends to zero, therefore (*γ* + *φ*)*I* → (1− q_1_)*αA*. Thus,

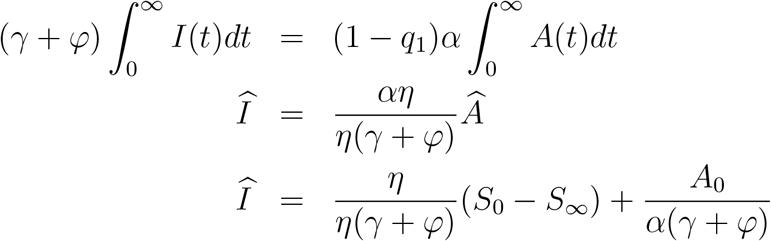

Let’s consider the first equation of our baseline model,

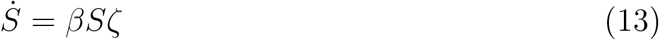

We could rewrite (13), as follows

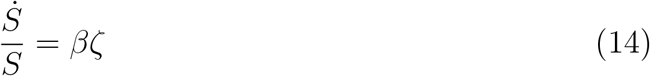

Integrating, from 0 to infinity, the left hand side, we obtain

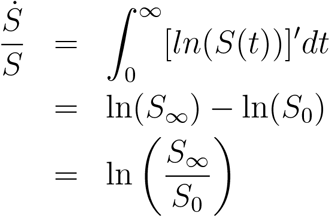

Now let’s integrating, from 0 to infinity, the right hand side, we assume (11) to have a constant denominator and then only replace each integral by *Ê, Â*, and *Î*.

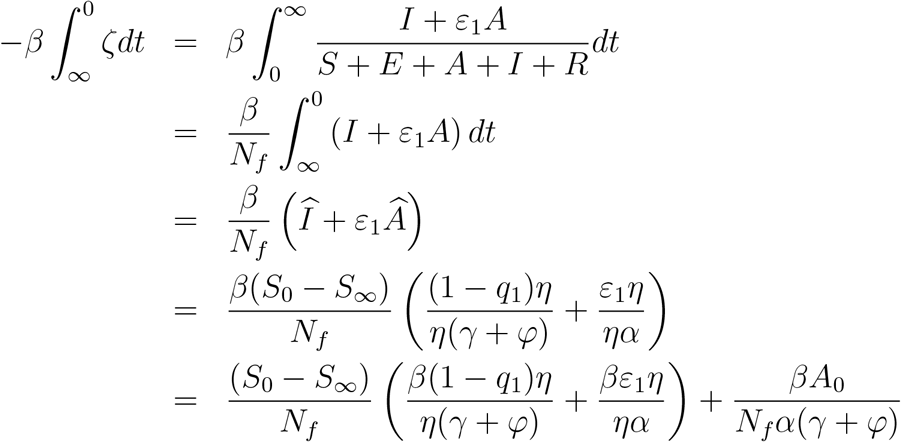

Where, we could substitute

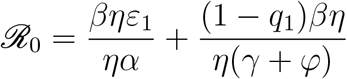

Therefore, substituting *N*_*f*_ according to (11). Also, considering

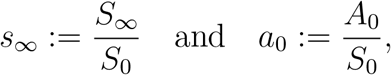

we obtain

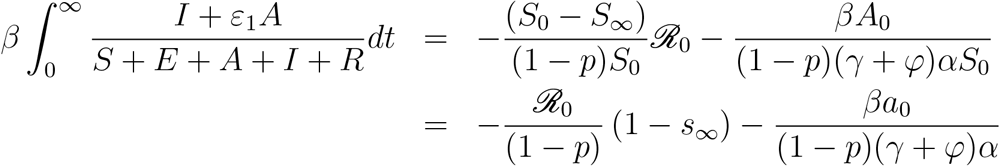

Then, the final epidemic size will be given implicitly by

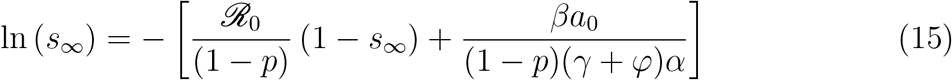

Let’s define

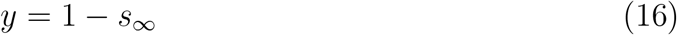

Therefore, by (16), the expression (15) is equivalent to

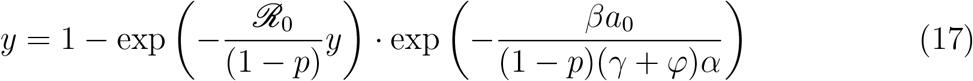

Therefore, by numerical approximations (taking *q*_3_ = 0.5, *β* = 0.6237, *A*_0_ = 1, and the other parameters as in Table 2) we could obtain that the final epidemic size will be *s* _∞_ = 10^−6^, i.e. S_∞_ = 10^−6^ · *S*_0_.

## D Appendix

### D.1 Uncertainty in parameter estimates

### D.2 Results Exponential Growth Fitting

### D.3 Results Generalized Growth Fitting

## E Epidemiological Results

### E.1 Compartmental COVID-19 Model Results

Figure 17 shows the results obtained after simulating the model presented in 2.2.2 with the parameters described in 2 and the fraction of population sent into quarantine *ρ* = 0.1 besides considering a limited number of ICU beds according to the data corresponding to each province.

**Figure 17:**
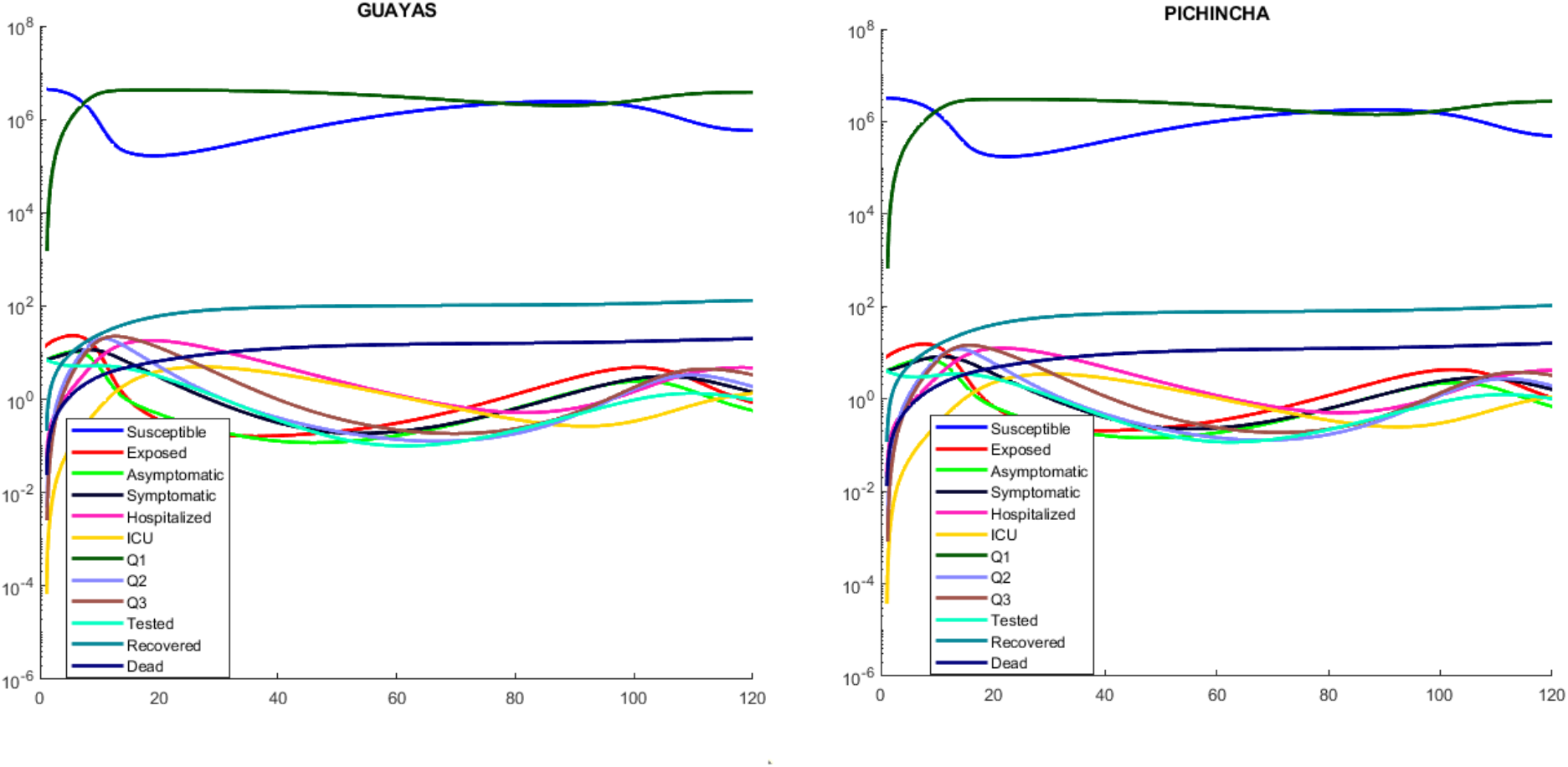
Simulation of the complete model without mobility (Model 2.2.2), considering limited number of ICU beds for two of the most affected provinces, for two of the most affected provinces

